# Fine grained two-dimensional cursor control with epidural minimally invasive brain-computer interface

**DOI:** 10.1101/2025.10.06.25337264

**Authors:** Ruwei Yao, Wenjianlong Zhou, Dingkun Liu, Wenzheng Li, Fangshuo Liang, Tao Liu, Honglai Xu, Wang Jia, Bo Hong

**Affiliations:** Department of Biomedical Engineering, School of Medicine, Tsinghua University; Department of Neurosurgery, Beijing Tiantan Hospital, Capital Medical University; Neuracle Technology Co., Shanghai, China

## Abstract

Brain–computer interface (BCI) can assist paralyzed patients in controlling external devices and improve their quality of life. However, existing intracortical BCIs entail risks of infection and challenges of long-term stability. In this study, we report on a tetraplegic patient implanted with our newly developed wireless minimally invasive BCI, NEO, in which eight Pt-Ir electrodes were placed epidurally over the hand area of the right sensorimotor cortex to record field potentials. We found that epidural neural signals from the hand area simultaneously represented both contralateral and ipsilateral movements. The spatio-spectral patterns of different movements exhibited prominent distinctions, revealing a bilateral representation structure of limb movements. Moreover, when two limb effectors (e.g., hand and elbow) moved simultaneously, their neural patterns exhibited non-additive changes relative to single movements. Based on these findings, we proposed a fine grained bilateral single/dual-movement decoding scheme for two-dimensional target control, thereby extending the degrees of freedom (DoF) of minimally invasive BCI systems and enhancing the information transfer rate (ITR). In two-dimensional center-out and web-grid tasks, the system achieved mean Fitts’ ITRs of 36.7 bpm and 30.0 bpm, respectively, with hit rates exceeding 91%. Neural recordings remained stable for over 18 months, and the decoder maintained stable performance for over 6 months without recalibration, demonstrating the safety and reliability of long-term home use.

## Introduction

Spinal cord injury interrupts the transmission of cortical neural signals to the limbs, leaving patients with nearly permanent tetraplegia and motor impairment^1^. Motor brain-computer interfaces establish a direct digital connection between the human brain and external devices such as computers by decoding neural signals^2,3^. Non-invasive neural recordings, such as electroencephalogram (EEG), typically have a relatively low signal-to-noise ratio^4,5^ and are commonly employed to discriminate among a finite repertoire of movements and the resting state. In contrast, implanted BCIs (iBCIs) provide neural recordings with superior spatiotemporal resolution. Implanted electrode arrays (MEAs) or microwire electrodes enable the recording of single-unit activity and have been used to control robotic arms^4-8^, computer cursors^9-11^, or to restore partial motor function through electrical stimulation^12,13^.

However, intracortical single-neuron recording BCIs have been found to cause tissue damage and scarring, while also facing potential electrode displacement, which in turn leads to degradation in recording quality^14-17^. Increasingly, research efforts have focused on balancing bandwidth with safety and stability in BCI systems, such as electrocorticography (ECoG)-based BCI^18^. ECoG enables the recording of intracranial field potentials from either subdural or epidural locations without penetrating the cortex, offering relatively high spatial resolution, robust noise immunity, and long-term stability^18-20^. Many intraoperative and short-term ECoG implantation studies have already demonstrated its potential for decoding different limb movements^21^, hand gestures^22^, and related motor functions. Nevertheless, the long-term goal of BCI systems development is to move from neurosurgical explorations to sustainable home use. To achieve this, several challenges remain, including the open cranial wounds necessitated by wired signal transmission^23^, as well as the risks of neuronal damage and infection associated with large-scale electrode implantation^23^, and long-term signal instability issues caused by electrode displacement^24^, among others.

Another major challenge lies in neural decoding and the control degrees of freedom. In endogenous motor BCIs, researchers can link the attempted movements to desired BCI control signals (e.g., cursor movement directions^21,25-29^) via an arbitrary mapping paradigm^19^,. This approach, however, depends on incorporating a sufficient number of distinguishable movements into the mapping protocol^30^. Voluntary movement is generally considered to be contralaterally dominated, yet ipsilateral activation and representation has been observed in single unit recordings^31-33^, fMRI^34^, and ECoG studies^35^; EEG has also revealed ipsilateral alpha- and beta-band ERD^36^. These findings support the feasibility of decoding ipsilateral movements^31^ and gestures^37^, as well as using ipsilateral activity to control one-dimensional cursor movements^38^. Another strategy introduces movements of multiple effectors (e.g., hand, elbow) into the encoding-mapping paradigm. Demonstrations of bimanual control^39,40^ and full-body exoskeleton control^41^ suggest that multi-effector schemes can expand the degrees of freedom available to BCIs. However, this requires dedicated decoding algorithms to address the nonlinear neural tuning changes associated with dual movements^42,43^. This study aims to integrate the aforementioned two strategies by constructing a bilateral and multi-effector neural encoding methodology based on unilateral epidural field potential recordings, thereby achieving multi-DoF target control and high information transfer rates in minimally invasive brain-computer interfaces.

Our recent study has developed the Neural Electronic Opportunity (NEO), a novel wireless minimally invasive epidural BCI^44^. The electrodes were implanted epidurally over the sensorimotor cortex, employing a wireless solution for power supply and data communication to minimize invasive damage and infection risks. We have validated its ability to achieve high-accuracy, real-time control of external devices, assisting paralyzed patients in grasping tasks and facilitating upper-limb functional rehabilitation^45^.

In this study, we report a brain-controlled two-dimensional target system for paralyzed patients based on NEO, enabling high information transfer rate (ITR) BCI target control. We analyzed motor neural encoding from epidural ECoG (eECoG) signals in the sensorimotor cortex, covering attempted movements of the contralateral and ipsilateral hand and elbow, as well as the simultaneous movements of two effectors (dual movements). Neural activity corresponding to different movements was separable in the spatio-spectral feature space, with axes approximately distinguishing the effectors and the laterality. This organization supports movement decoding and independent control across degrees of freedom. We further found that the neural encoding of dual movements is non-additive, highlighting the importance of incorporating dual movements into the mapping protocol with specialized decoding. Building on this, we proposed a bilateral single/dual-movement two-dimensional target control scheme, where movement decoding probabilities were weighted and mapped onto two-dimensional directions, and achieved online target control in both center-out and web-grid tasks. Owing to the stability of epidural signals, this two-dimensional BCI control system operates without requiring frequent recalibration and maintains long-term performance stability.

### Wireless minimally invasive brain-computer interface

Our team designed an implantable minimally invasive wireless brain-computer interface system, NEO, to achieve safe and reliable neural signal recording and long-term effective decoding. The NEO implant consists of a processor, an internal coil, and an epidural electrode. The implantable processor is encapsulated in a titanium case, measuring approximately 25 mm in diameter and only as thick as two stacked coins (Fig. 1a). It is embedded into the cranial surface via a 2-mm bone recess (Fig. 1b). Two sets of Pt-Ir electrodes (each set containing four electrodes, 3.2 mm in diameter, with 8 mm center-to-center spacing) were placed epidurally over the right hemisphere primary motor cortex (channels 1-4) and primary somatosensory cortex (channels 5-8), respectively (Fig. 1c), and connected to the processor through lead wires routed via micro-tunnels drilled through the skull.

**Figure 1.**
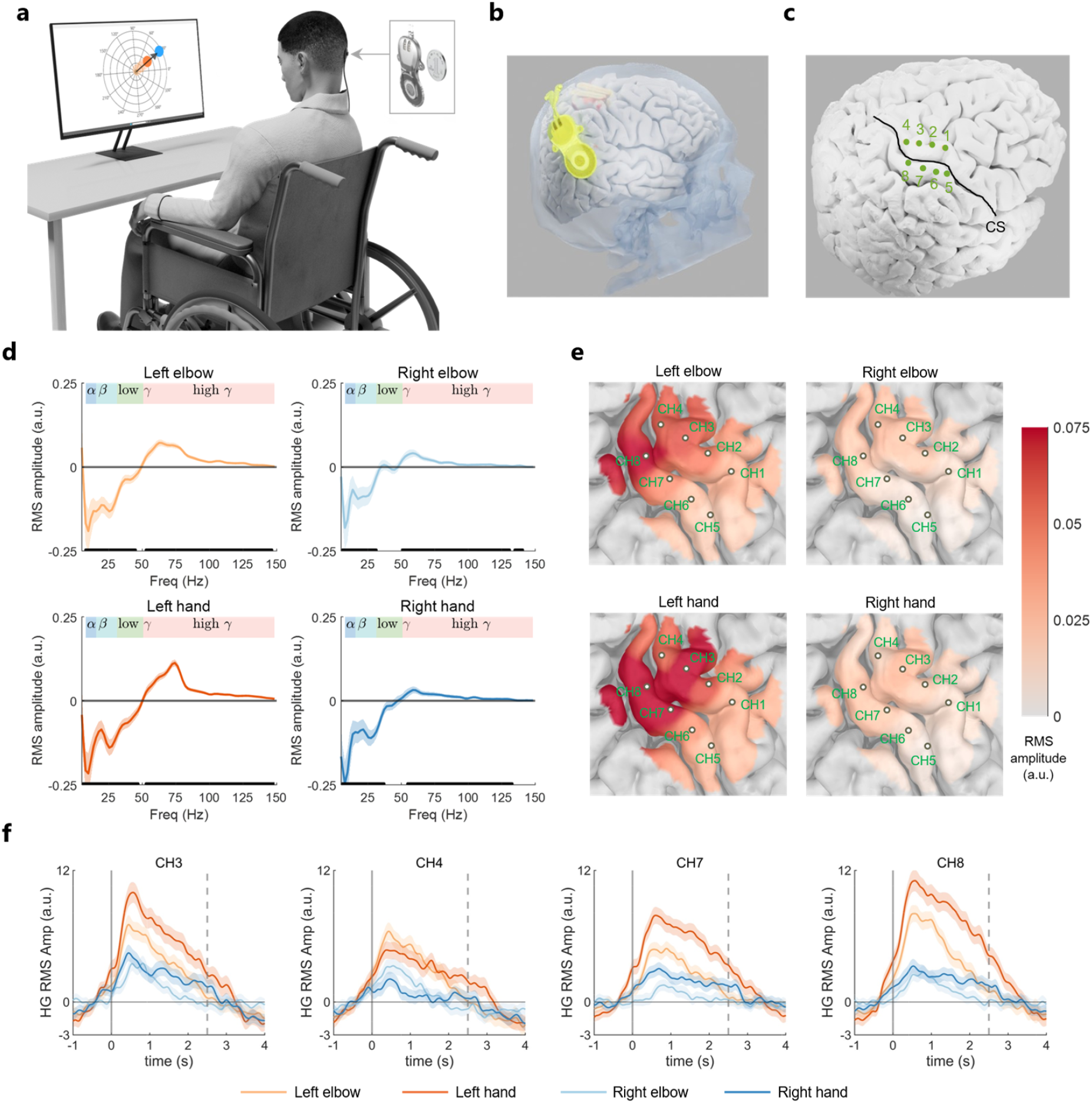
Wireless minimally invasive epidural BCI for recording motor-related neural activity. **a**. The NEO implant consists of a processor, an implanted coil, and epidural electrodes. An external coil, attached to the scalp, couples with the implanted coil to wirelessly transmit signals and deliver power. The NEO enables paralyzed patients to control a two-dimensional target. **b**. The implant is embedded on the outer surface of the skull and connected to epidural electrodes through a micro-tunneled transcranial wire lead. **c**. Two set of 4-electrode arrays are placed epidurally over the hand area of the sensorimotor cortex. **d**. Frequency-domain tuning of a representative electrode (CH2) in the primary motor cortex. The electrode is tuned to movements of both contralateral and ipsilateral effectors (elbow, hand; n = 380 trials for each movement). Each subpanel shows event-related RMS amplitude changes across different frequency bands (α: 8–12 Hz, β: 14–30 Hz, low γ: 30–50 Hz, high γ: >50 Hz) from 0–150 Hz. The task phase was defined as 200–1200 ms, with −1000–0 ms serving as baseline. Shaded areas indicate standard error. Thick black bars mark frequency bands with statistically significant deviations from baseline (one-sample t test, *p* < 1e−3). **e**. Tuning heatmaps for each movement across the sensorimotor cortex. Color scale indicates the average RMS amplitude of the high γ band (55–95 Hz) during 0–2 s after movement onset, relative to the −1–0 s baseline, interpolated across electrodes. **f**. Time course of high γ band (55–95 Hz) RMS amplitude for different movements recorded from a representative electrode in the sensorimotor cortex. Shaded areas indicate standard error. Neural activity was convolved with a Gaussian smoothing kernel (σ = 300 ms) for denoising.

The NEO implant does not contain a battery. Instead, it adopts a wireless communication and power supply solution. The implanted coil is inductively coupled to an external coil positioned on the scalp, which is held in place by a magnet. Through this inductive link, the implanted coil receives power from the external device. Simultaneously, it wirelessly transmits eight-channel epidural ECoG signals (sampled at 1000 Hz) to the external device, which then relays the data to a computer. The BCI decoding algorithm processes these neural signals to generate predictions of different movement intentions, enabling real-time online control of interactive devices such as a computer screen target (Fig. 1a).

### Neurosurgical implantation

This study was part of the clinical trial of the implantable closed-loop brain-computer interface system (NEO) (clinicaltrials.gov; NCT05920174), and was approved by the Ethics Committee of Tiantan Hospital in August 2023. We recruited a male patient who had suffered a complete (C4) spinal cord injury (AIS-A, Extended Data Fig. 1) due to an accident. The patient had limited upper limb motor function and was unable to move the upper arm limbs independently or perform any voluntary hand movements except for limited flexion of the left elbow.

To ensure accurate electrode placement, we conducted a functional MRI (fMRI) study to identify cortical regions most responsive to hand-related sensorimotor activity. Both passive and active movement paradigms were used to localize the sensorimotor areas associated with the patient’s hand, as well as the cortical regions corresponding to residual ascending sensory signals (Extended Data Fig. 2a,b). A T1-weighted MRI scan was acquired to reconstruct cortical anatomy, and virtual electrode placement in 3D visualization software demonstrated coverage of the principal activation regions (Extended Data Fig. 2c). For implantation planning of the internal unit, cranial bone thickness at the candidate sites was evaluated to ensure adequate support for embedding the device. Based on the patient’s CT images, cranial thickness was computed (Extended Data Fig. 2d), and the implant position was adjusted in 3D visualization software to confirm a suitable site. Finally, the planned locations of the internal device and electrodes were exported to the surgical navigation system to guide implantation.

The minimally invasive BCI implantation surgery was successfully completed at Beijing Tiantan Hospital. The patient’s condition remained stable and recovered well after surgery. Postoperative CT images were acquired (Extended Data Fig. 2e,f) to extract electrode locations, which were then co-registered with the preoperative MRI. Four electrodes (CH1-4) were located above the precentral gyrus, and four electrodes (CH5-8) were above the postcentral gyrus, covering the regions showing the strongest hand-related activation during fMRI motor imagery. Among them, CH1-3 covered Brodmann area 6d, CH4 covered Brodmann area 4, and CH5-8 covered Brodmann area 1-3.

### Neural encoding of ipsilateral and contralateral movements

Using the NEO BCI, we investigated the neural representations of epidural field potentials in the sensorimotor cortex during different upper-limb movements (contralateral and ipsilateral elbow flexion and hand grasp). In the experiments, the patient was required to attempt to perform corresponding limb movements according to the text or picture prompts on the computer screen. Movement-related neural responses were reflected in relative power changes across different frequency bands. We conducted time-frequency analysis on multi-channel neural signals and observed significant limb movement-related event-related spectral perturbation (ERSP) (Extended Data Fig. 3b). Relative to the preparation phase (baseline), the onset of movement was accompanied by typical event-related synchronization (ERS) in the high-gamma frequency band (>50Hz), indicative of increased local neuronal firing in the sensorimotor cortex. Event-related desynchronization (ERD) occurs in the low frequency bands covering alpha (8-12Hz), beta (14-30Hz), and low gamma (30-50Hz). At lower frequencies, power increases in the delta band (1-4 Hz) were observed during movement planning, execution, and return phases, potentially reflecting slow cortical potentials, also known as movement-related cortical potentials (MRCPs). Following movement termination, high-gamma power returned to baseline levels, while the beta band exhibited a pronounced post-movement rebound.

We investigated the differences in neural activity among different movements. We calculated the root mean square (RMS) amplitude features of different frequency bands to evaluate neural representations, and observed significant differences in the responses of multiple electrodes to various movements such as elbow flexion and hand grasping (see Fig. 1d for frequency-domain energy tuning of an example electrode, CH2). We found that the unilateral epidural field potentials could represent not only contralateral (left) movements relative to the implant site but also ipsilateral (right) movements. In both the low-frequency range (3-25 Hz) and the high-gamma band (55-125 Hz), the RMS amplitude differences between movement and baseline were statistically significant (one-sample t-test, *p* < 1 × 10^−3^). For different movements, distinct suppression peaks were observed in both alpha and beta bands, with relatively high trial-to-trial variability and comparable suppression amplitudes across movements. By contrast, the high-gamma band showed clear distinctions between movement types (Fig. 1e-f): contralateral movements elicited stronger ERS than ipsilateral ones. Anatomically, high-gamma activity during elbow movement was more concentrated dorsally over BA4 and BA1-3 (CH4, CH3, CH8), whereas hand movements were represented more ventrally (CH2, CH3, CH7, CH8). These findings are consistent with classical sensorimotor cortical somatotopic maps.

Considering the task-specific differences in high-gamma activity, we further analyzed the distinctions structure of neural representations across different movements. For the task period (200-1200 ms), multichannel, multiband RMS amplitudes (covering 50-150 Hz) were subjected to principal component analysis (PCA; see Methods) to identify two neural dimensions for visualizing movement differences. Neural patterns are projected onto the first two principal components (Fig. 2a). During isolated single-effector movements, contralateral elbow flexion, contralateral hand grasps, and ipsilateral movements formed clearly separable clusters. Furthermore, the first dimension effectively separated contralateral and ipsilateral movements, while the second dimension approximately distinguished between two different effectors: the elbow and the hand. This appears to indicate the existence of compositional coding^33^ at field potentials level.

**Figure 2.**
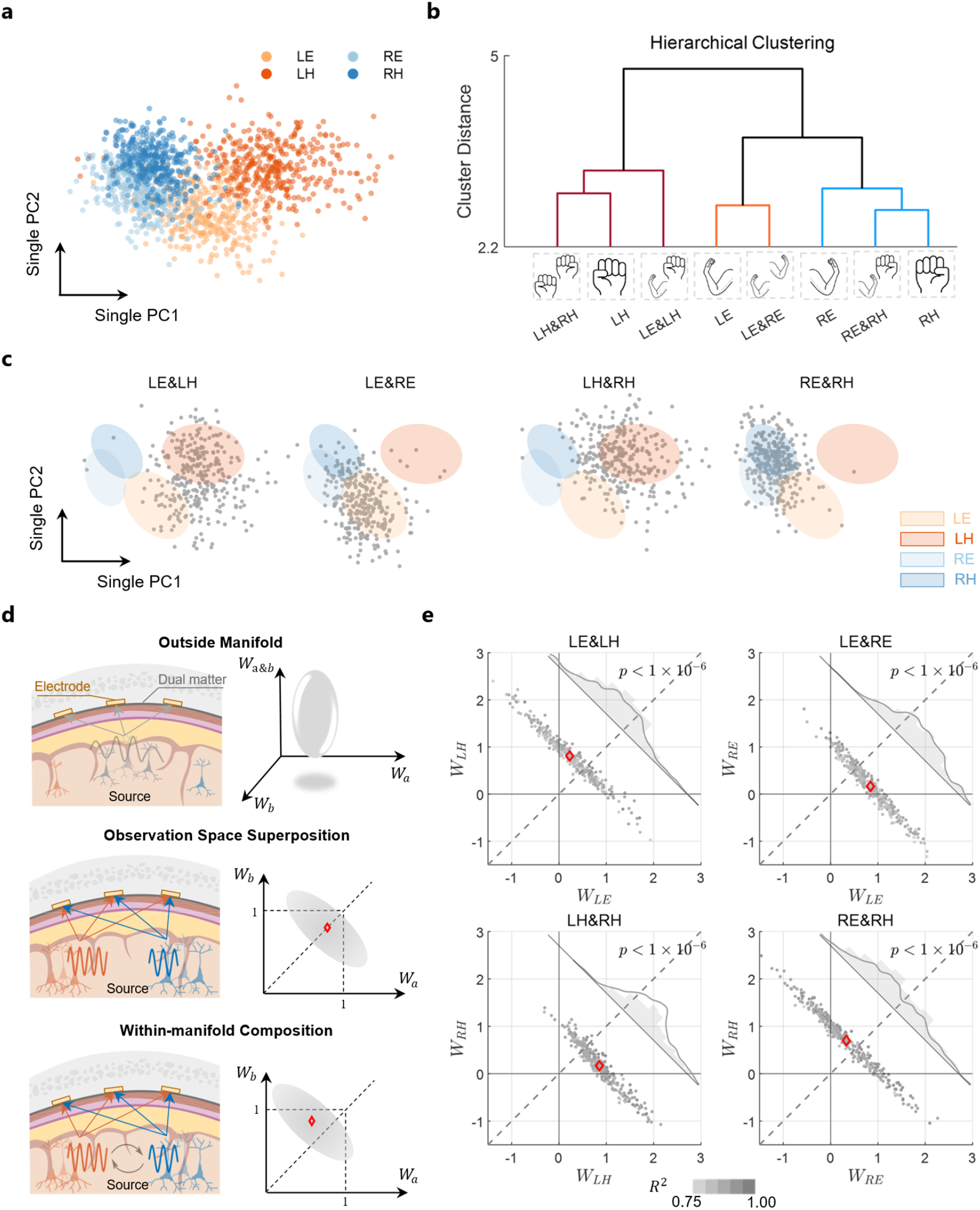
Bilateral single and dual movement neural representational structure. **a**. High-gamma (HG) spatiotemporal patterns of single movements (LE: left elbow, LH: left hand, RE: right elbow, RH: right hand) projected onto the first two principal components of neural activity. Each dot represents one trial. **b**. Hierarchical clustering (Ward’s criterion) of pairwise Mahalanobis distances between neural centroids, revealing a representational structure described by the dendrogram. **c**. Low-dimensional projections of neural patterns for four dual movements onto the axes defined by the first two single-movement components. Each dot represents one dual-movement trial; ellipses indicate the 50% confidence intervals (CI) of the corresponding single-movement distributions. **d**. Alternative hypotheses for dual-movement neural encoding mechanisms. The *outside-manifold* hypothesis predicts the existence of independent neural dimensions or populations in the sensorimotor cortex specialized for dual movements, such that single movement bases explain only limited dual movement variance. The *observation-space superposition* hypothesis predicts that dual movements are merely the sensor-level superposition of single-movement activity, equivalent to a 1:1 additive projection. The *within-manifold composition* hypothesis predicts that dual movements are encoded by the same neural populations as the corresponding single movements but with non-additive interactions. **e**. Distribution of regression coefficients when dual movements were fitted by their corresponding two single movement component-bases (showing trials with R^2^ > 75%; outlier trials are retained in the plot). Diamonds mark the mean dual movement patterns in the *component-basis* space. The histogram quantifies distribution of distances of scatter points to the 1:1 symmetry axis (proportional to the difference between regression coefficients on the two bases).

### Two effectors move simultaneously

Based on the encoding analysis of the isolated movement of one effector (single movements), we explored the neural representation when two effectors move simultaneously (dual movements). Previous studies have demonstrated that dual movement combinations have unique encoding properties and are distinct from single movements^33,43^, thereby offering additional control degrees of freedom for BCI applications. However, the extent of potential interference when integrating both single and dual movement into an eECoG based BCI protocol remains unclear. Here, we sought to explore the independence between dual movements neural representations and their corresponding single movements, thereby providing prior insights for the design of BCI paradigms that integrate both single and dual movements.

We studied four basic unilateral single movements and their four corresponding dual movements, yielding eight conditions in total. The trial-averaged time-frequency patterns of multichannel RMS amplitude under eight conditions exhibited discernible differences (Extended Data Fig. 4a). To quantify the relationships among different movements in neural space, we measured Mahalanobis distances between the multidimensional spatio-spectral patterns (Extended Data Fig. 4b). Pairwise Mahalanobis distance matrices were computed between the mean activity patterns (neural centroids) of all movements (Extended Data Fig. 4c), and hierarchical clustering based on these distances revealed a structured representation of unilateral single and dual movements (Fig. 2b). First, a prominent axis clearly separated movements of different effectors (hand vs. elbow), with contralateral effectors showing stronger tuning and ipsilateral effectors weaker tuning. Second, contralateral effector representations distinguished hand and elbow movements with clear spatio-spectral differences. Finally, we observed that the pattern associated with simultaneous elbow-hand dual movements lay between those of the corresponding single movements, but was closer to that of the hand.

Since dual movements involve the simultaneous control of two effectors within the sensorimotor cortex, we anticipated that their neural patterns would more closely resemble those of the corresponding single movements. To test this hypothesis, the dimensionality reduction analysis described in the previous section was applied, projecting the four dual movements onto the two neural dimensions derived exclusively from single movements. It is shown that dual movements were projected between the clusters of their corresponding single movements (Fig. 2c). Such separability is critical for online motor decoding; otherwise, dual movement episodes occurring during transitions between two single movements could lead to discontinuous decoding jumps.

Previous studies based on neuronal spiking recordings in the primary motor cortex have revealed unique neural encoding characteristics and underlying mechanisms for bimanual actions^33,43^. Here, at the epidural level of the sensorimotor cortex, we extended the concept of population coding and treated the joint spectral-channel patterns in the high-gamma (HG) band as neural activity state^46^. This allowed us to analyze the relationship between the subspaces of single and dual movements. We proposed three alternative hypotheses regarding the neural representation of dual movements (Fig. 2d). The first is the *Outside-manifold* hypothesis, which posits that dual movements are encoded in a neural subspace very different from that of the corresponding single movements. This would imply that the sensorimotor cortex provides an entirely new representation for dual movements, such that only a small portion of their variance could be explained by single-movement neural dimensions. The second is the *Observation-space superposition* hypothesis, which suggests that dual movements simply reflect the simultaneous but independent neural activity of two single-movement populations, and that signals are directly superimposed in the sensor space; under this view, single and dual movements would share exactly the same subspace. The third is the *Within-manifold composition* hypothesis, which posits that single and dual movements are encoded largely within the same subspace, but not as a direct one-to-one superposition; instead, their representations exhibit mutual interactions. This is based on the consideration that natural motor behavior typically requires the cooperation of multiple effectors, the sensorimotor cortex may coordinate the neural encoding of multi-movement actions.

To test this, we constructed three types of neural “bases” to linearly regress the neural patterns of various dual movements, with each basis consisting of two distinct vectors. The first was the *self-basis*, defined as the first two principal components of the dual movement itself, representing its intrinsic neural subspace. The second was the *orthogonal-basis*, defined as the last two principal components of the same dual movement. As these dimensions are expected to explain little variance, they serve as a surrogate for the outside-manifold scenario. The third was the *component-basis*, constructed from the mean patterns of the two corresponding single movements, representing the subspace spanned by those two constituent single movements.

We quantitatively evaluated the goodness-of-fit of linear regression using the three types of bases for the four dual movements (Extended Data Fig. 5). As expected, the *self-basis* consistently yielded the best fits across all dual movements. For all cases, the *component-basis*, constructed from the corresponding single movements, also achieved comparably high goodness-of-fit, showing no statistically significant difference from the *self-basis* (two-sample *t*-test, *p* > 0.1, except *p* = 4 × 10^−3^ for RE & RH), but significantly greater than the *orthogonal-basis* (two-sample *t*-test, *p* = 2 × 10^−7^ for LE & LH, *p* = 0 for LE & RE, *p* = 6 × 10^−6^ for LH & RH, *p* = 4 × 10^−5^ for RE & RH). In addition, we introduced a *non-component-basis*, constructed from non-corresponding single movements (i.e., entirely different effectors or contralateral movements; for example, LE & LH are non-corresponding to RE and RH, and LH & RH are non-corresponding to LE and RE), and compared its performance. Across all dual movements, the *component-basis* again achieved significantly better fits than the *non-component-basis* (two-sample *t*-test, *p* = 0 for LE & LH, *p* = 3 × 10^−5^ for LE & RE, *p* = 6 × 10^−3^ for LH & RH, *p* = 2 × 10^−7^ for RE & RH). Taken together, these results suggest that dual movements and their corresponding single movements are largely encoded within the same spatio-spectral subspace, which conforms to the *Within-manifold composition* hypothesis.

Since the patterns were obtained in the eECoG sensor space, we asked whether the result of the shared subspace between single and dual movements was merely due to the volume conductor effect (*Observation Space superposition*). We constructed a surrogate dual movement neural dataset under the superposition hypothesis (see Methods), and performed the same regression analysis on the empirical dual movement neural data and the surrogate data. For the empirical data, Fig. 2e shows the regression coefficients of the two *component-basis*. We found that: first, the regression coefficients were not clustered around the point [1, 1], but rather aligned along the line *w*_*a*_ + *w*_*b*_ = 1, indicating that the HG energy of the dual movement signal was “normalized”, and the total signal energy did not superpose due to the combination of more movements. Second, the ratio of the regression coefficients was not distributed near 1:1 (on the gray dashed line). For purely unilateral dual movements, the regression coefficients for the hand *component-basis* were significantly greater than those for the elbow *component-basis* (Wilcoxon signed-rank test, *p* = 5.2 × 10^−18^ for LE&LH, mean coefficient ratio *w*_*LE*_: *w*_*LH*_ = 0.27; *p* = 3.9 × 10^−7^ for RE&RH, mean coefficient ratio *w*_*RE*_: *w*_*RH*_ = 0.47), indicating that the hand, relative to the elbow, acted as the dominant effector. In contrast, for purely single-effector dual movements, the contralateral *component-basis* overwhelmingly dominated over the ipsilateral *component-basis* (Wilcoxon signed-rank test, *p* = 5.6 × 10^−22^ for LE&RE, mean coefficient ratio *w*_*LE*_: *w*_*RE*_ = 5.15; *p* = 3.9 × 10^−31^ for LH&RH, mean coefficient ratio *w*_*LH*_: *w*_*RH*_ = 5.06). This effect was generally consistent across channels (Extended Data Fig. 5c). By contrast, for the simulated surrogate data under the *Observation Space Superposition* hypothesis (Extended Data Fig. 5b), no significant differences were observed in coefficient (Wilcoxon signed-rank test, *p* > 1 × 10^−3^).

Overall, these results demonstrate that at the epidural field-potential level of the sensorimotor cortex, the encoding of bilateral single movements and dual movements is better explained by the *Within-manifold composition* hypothesis, consistent with observations from single unit recordings^43^. From a BCI decoding perspective, our findings suggest that treating dual movements as independent classes may be more advantageous for neural control.

### Two-dimensional control scheme for bilateral single-dual movement

The analysis of neural representations for different movements inspired the design of BCI paradigms and decoding algorithms that enable independent multidimensional control with high information transfer rates. We developed a closed-loop BCI system that allows a tetraplegic patient to continuously control two-dimensional targets through movement intentions. Specifically, we proposed a two-dimensional control scheme that integrates both bilateral single movements and dual movements (see Methods), with the controllable repertoire comprising four single movements and their four corresponding dual movements. The scheme involves two steps: first, we trained a classifier to discriminate among the eight movement intentions as well as rest; second, a fine-grained mapping vectors set projected the posterior classification probabilities into two-dimensional velocity vectors, thereby establishing a mapping from movement intentions to target velocity (Fig. 3a).

**Figure 3.**
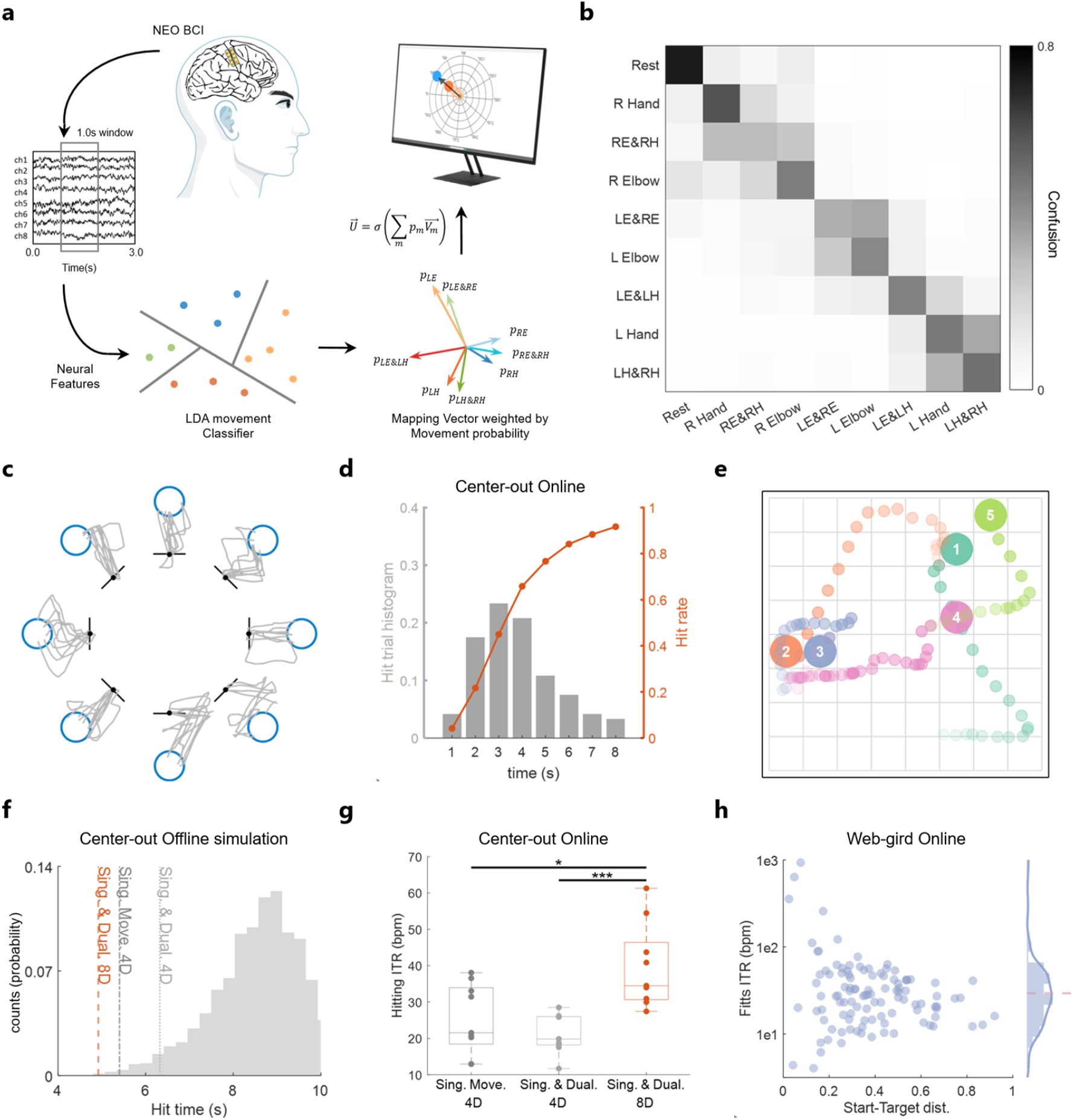
Brain-controlled two-dimensional target tasks. **a**. Schematic of closed-loop BCI two-dimensional control. The user controls an orange target to hit blue targets on the screen. Multichannel neural data within a 1000 ms window were transformed into log-power spectral density features (0–100 Hz). A linear discriminant classifier estimated probabilities of belonging to eight movement classes and rest. The velocity direction was determined by the probability-weighted sum of mapping vectors, while velocity magnitude was controlled by a gating function. **b**. Normalized confusion matrix of bilateral single/dual movement and rest classification, obtained from 100 iterations of 10-fold cross-validation. **c**. Controlled target trajectories (gray lines) toward cued targets (blue circles) in online Center-out task experiments, aggregated across all successful trials from five testing sessions. Start positions (black dots) were shifted outward to avoid trajectory overlap in visualization. **d**. Histogram of hit times and cumulative hit rates for the Center-out task using the Sing.&Dual 8D mapping scheme. **e**. Illustration of online web-grid control trajectories. Controlled target paths are shown for five sequentially presented cued targets (order indicated by numbers), with trajectory transparency increasing over time. **f**. Offline Center-out task simulations with surrogate mapping vector sets. Histogram of average hit times, with vertical lines marking mean hit times for three representative mapping schemes. **g**. Online Center-out task results comparing different mapping vector schemes. Each dot represents the Hitting ITR computed from one full cycle of eight targets acquisitions. **h**. Online web-grid testing (Month 1, five sessions) showing Fitts’ ITR across hit trials. The x-axis indicates distance from start to cued target. The average Fitts’ ITR reached 60.05 bpm (pink dashed line). Because nearby targets yielded inflated ITR values, excluding outliers or using the median provides a more robust estimate.

We first employed a linear discriminant classifier to perform multiclass classification of the spatio-spectral patterns of eECoG signals, yielding an offline confusion matrix across the eight movement conditions and rest (Fig. 3b), achieving a macro-accuracy of 0.48 ± 0.028 SD and an F1-score of 0.47 ± 0.026 SD (averaged over 100 iterations of 10-fold cross-validation), with the chance level limited at 11.1%. All movement patterns were highly distinguishable from rest (classification accuracy for rest: 89.6% ± 3.1% SD). Consistent with the clustering analysis, three clusters (left elbow, left hand, and right side) were clearly separated. Misclassifications mainly occurred within clusters; nevertheless, intra-cluster discrimination remained significantly above chance level.

For the mapping rules, we sought to design velocity control such that: (i) the theoretical precision of two-dimensional target direction control is maximized (corresponding to higher information transfer rates); (ii) the rest state corresponds to near-zero velocity; and (iii) align with user intuition; for instance, the direction corresponding to the dual movement A&B should approximately lie along the angular bisector of the directions associated with movements A and B. To this end, we established a two-dimensional BCI channel model within the information-theoretic framework, with the optimization objective of maximizing the BCI information transfer rate (see Methods). This yielded a fine-grained set of eight directional mapping vectors consistent with the neural representational structure (Extended Data Fig. 7a), referred to as the Sing.&Dual 8D scheme. Two-dimensional target velocity was then obtained by weighting the mapping vectors with the decoding posterior probabilities, analogous to the classical population vector method^47^, with velocity magnitude gated by the posterior probability of the rest state.

### Two-Dimensional Control Test

We asked whether the NEO bilateral single/dual movement scheme could support continuous two-dimensional control. The patient used the NEO system to perform a center-out task (Extended Data Fig. 6a), in which a brain-controlled target was driven to touch one of eight peripheral targets on the screen as cued (cued target). Each trial allowed up to 8 s for a successful hit, after which the task advanced either upon target acquisition or timeout. Fig. 3c shows representative trajectories of hit trials across five test sessions. The efficiency and robustness of the brain-controlled trajectories were evaluated, with an average trajectory distance ratio of 2.17 ± 0.99 SD, trajectory error of 0.27 ± 0.15 SD, and trajectory variability of 0.19 ± 0.10 SD. The decoding algorithm achieved 91.67% success within 8 s (84.17% within 6 s), with an average hit time of 3.28 ± 1.67 s, corresponding to a mean Fitts’ information transfer rate (ITR) of 36.74 ± 22.74 bpm (Fig. 3d).

To further test more naturalistic target control, the patient performed a web-grid task (Extended Data Fig. 6b), in which the target was continuously brain-controlled (without resetting to the center) to reach randomly appearing targets in an 8 × 8 grid within 14 s. After training, the participant was able to complete the task proficiently (Fig. 3e), achieving an average hit rate of 90%. The mean hit time was 4.64 ± 3.31 s, yielding a mean Fitts’ ITR of 29.88 ± 18.23 bpm (after excluding upper adjacent outliers), with a median Fitts’ ITR of 27.30 bpm (Fig. 3h).

We further explored, via theoretical simulations, whether the bilateral single/dual movement (Sing.&Dual 8D) scheme is efficient, and whether the inclusion of dual movements and the dimensionality of the mapping vector set are critical. First, we conducted virtual offline simulations for center-out task (see Methods) to validate our theoretically optimized results, constructing scrambled surrogate mapping vector sets as controls. The Sing.&Dual 8D paradigm yielded shorter average target acquisition times in the simulated center-out task than all surrogate schemes (Fig. 3f). We also evaluated two additional representative velocity control schemes (Extended Data Fig. 7a): (i) Sing. Movement 4D: classification of only the four single movements (confusion matrix shown in Extended Data Fig. 7c) mapped to four preferred directions; (ii) Sing.&Dual 4D: the eight single and dual movements relabeled into four classes according to pattern similarity and mapped to four preferred directions (confusion matrix in Extended Data Fig. 7d). In the simulations, the Sing.&Dual 8D scheme consistently outperformed both the Sing. Movement 4D and Sing.&Dual 4D schemes (Fig. 3f).

Finally, we tested the Sing. Movement 4D and Sing.&Dual 4D schemes in actual online center-out tasks (Fig. 3g) and compared them with the Sing.&Dual 8D scheme. To jointly account for target acquisition time and success rate, we defined a Hitting ITR metric (see Methods). The Sing.&Dual 8D scheme achieved the highest Hitting ITR (39.69 bpm ± 11.64 SD). In comparison, schemes with fewer preferred directions—Sing. Movement 4D (25.30 bpm ± 9.70 SD) and Sing.&Dual 4D (21.43 bpm ± 5.49 SD)—performed worse, significantly lower than the Sing.&Dual 8D scheme (*p* = 0.032 for Sing. Movement 4D; *p* = 8e-5 for Sing.&Dual 4D), and exhibited reduced trajectory smoothness (Extended Data Fig. 7e,f). These results indicate that incorporating dual movements and designing an optimal mapping vector set significantly boosts the information transfer rate of two-dimensional BCI control.

### Long-term stability of the NEO system

Stable signal quality, implant safety, and decoding performance are critical prerequisites for long-term home use of BCIs. We performed long-term recordings with the NEO BCI implant to quantify the stability of neural signals and decoding algorithms. Over nearly 18 months post-implantation (from day 0 to ∼523 days), electrode contact impedance increased only during the first 3 months after implantation (averaged 0.012 *k*Ω/day, likely due to postoperative tissue healing) and remained stable thereafter without significant changes (Extended Data Fig. 8c).

We tracked the quality and features of resting-state signals over a long period. Both epidural neural signals and multi-channel power spectral densities remained stable and consistent over the nearly 17-month span (Extended Data Fig. 8a), with the RMS amplitude increasing at an average rate limited to 0.01 *μ*V/day. We calculated the temporal evolution of signal power across different frequency bands (Extended Data Fig. 8b). Linear regression indicated that the low-frequency band (0–50 Hz) exhibited a minor average increase, limited to 0.0042 dB/day, while high-gamma activity (50–200 Hz) showed no significant increase or decrease (rate of change < 1e-3 dB/day).

We further evaluated the robustness of offline classification for different movements. Without any recalibration, the bilateral single/dual movement classifier was tested for cross-time generalization. As shown in Fig. 4b, the classifier remained stable for over 190 days post-construction, achieving an average movement classification accuracy of 42.28% (Chance level=11.11%), with no significant decline in bilateral single/dual movement accuracy observed (correlation *r* = 0.054, linear regression *p* = 0.834, significance level *α* = 0.05). Confusion matrices spanning more than two months remained largely consistent (Fig. 4a), preserving clear separability among resting-state versus movement, contralateral elbow, contralateral hand, and ipsilateral movement clusters.

**Figure 4.**
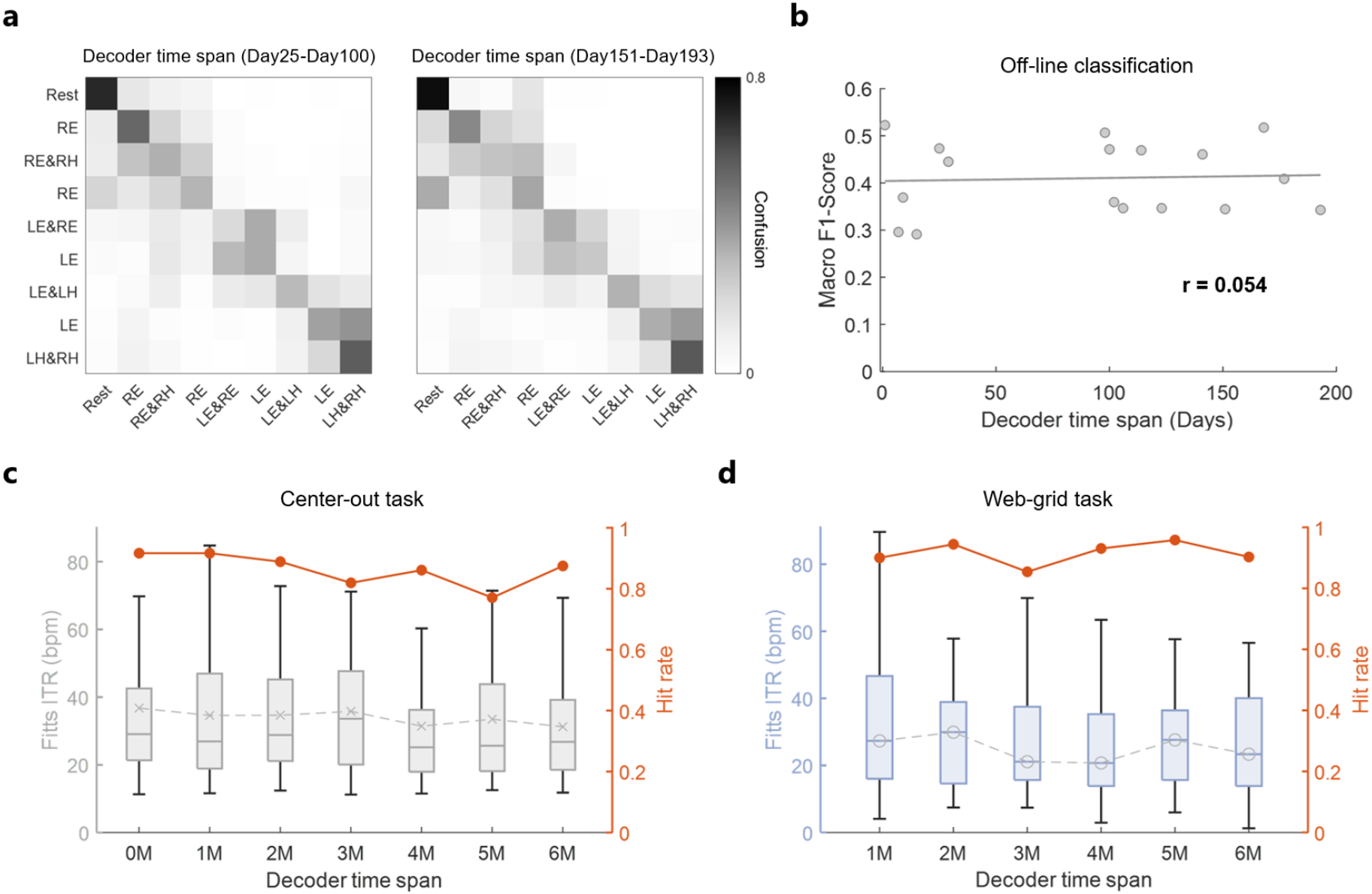
Long-term stability of two-dimensional control. **a**. Offline classification confusion matrices of bilateral single/dual movements and rest during long-term application. The classification results of the decoder for Day25-100 (left) and Day151-193 (right) after its construction are shown respectively. **b**. Macro F1-score of long-term generalization of offline classification of bilateral single/dual movements and rest. Each scatter point represents the average F1-score within the session on the corresponding date. **c**. Fitts’ ITR and hit rate of the online Center-out task with the decoder applied for several months. **d**. Fitts’ ITR and hit rate of the online web-grid task for several months.

Finally, we assessed the long-term stability of online BCI performance. Without recalibrating the trained two-dimensional decoding model and the control scheme, we conducted monthly tests starting from the training month. Across a 6-month span, the average Fitts’ ITR for the closed-loop center-out task was 33.99 bpm (Fig. 4c), with only a slight decline over time (−0.026 bpm/day, 95% CI: [−0.048, −0.005], *p* = 0.026, *α* = 0.05). The monthly average hit rate within 8 s was 86.41%, showing no significant decrease (*β* = −0.016%/day, 95% CI: [−0.048, 0.006], *p* = 0.12, *α* = 0.05). For the web-grid task, Fig. 4d illustrates performance over 1 to 6 months, where the average Fitts’ ITR remained at 24.99 bpm with no significant decline (β = −0.026 bpm/day, 95% CI: [−0.113, 0.061], *p* = 0.457, α = 0.05). The average hit rate within 14 s was 91.5%, which also showed no significant change over time (β = 0.004%/day, 95% CI: [−0.024, 0.031], *p* = 0.72, *α* = 0.05).

In summary, we demonstrate the long-term stability of both the device and neural signal characteristics of the wireless minimally invasive BCI. Combined with a robust bilateral single/dual movement decoding scheme, epidural signals can support high-ITR, long-term usable two-dimensional target control.

## Discussion

We utilized a wireless, minimally invasive epidural BCI to decode motor intentions from the sensorimotor cortex, enabling an individual with tetraplegia to voluntarily control two-dimensional target. The system has been implanted and operating for more than 18 months with stable performance. Two-dimensional control was achieved via a bilateral single/dual movement decoding and mapping scheme. Both theoretical optimization and online testing confirmed that this approach can achieve high information transfer rates. This scheme leverages the representational structure of contralateral and ipsilateral movements in the spatio-spectral feature space. Importantly, dedicated decoding of dual movements proved beneficial, as we verified that dual movement neural activity is not a simple linear superposition of single movements.

Implantable wired microelectrode arrays BCI have previously demonstrated the highest performance in target control and substantial degrees of freedom in robotic arm control (up to 10D)^48^. However, their long-term use has been limited by poor biocompatibility^49^, glial scarring^50-53^, electrode migration^53^, infection^23^, and high signal variability^24,54,^. Here, we report a wireless minimally invasive approach in which electrodes are secured epidurally, avoiding direct penetrating the dura, while the wireless and battery-free design minimizes infection and safety risks. Over 18 months, signal features remained stable; limited increases in low-frequency power did not impair the movements decoding, substantially reducing recalibration frequency and cost. Although demonstrated in a single patient with paralysis, the minimally invasive safety and robust cortical tuning suggests that this system and decoding framework could be extended to a broader population of individuals with SCI or ALS.

Our study demonstrates the potential of decoding contralateral, ipsilateral, and bilateral movements from unilateral eECoG recordings. By leveraging spatio-spectral high-gamma features of epidural field potentials, we revealed a prominent bilateral representational structure, where laterality and effector can be dissociated along distinct neural dimensions. These findings are consistent with previous studies^35,37,55,56^, and align with the compositional coding hypothesis previously proposed from microelectrode array recordings in paralyzed individuals—both at the level of neuron spike firing rates^33^ and intracortical local field potentials^57^. The compositional coding framework posits separable dimensions for laterality and for specific effectors^33^. Importantly, such a property supports the design of independently controllable BCI degrees of freedom. In our case, this enabled a user with unilateral implantation to intuitively and continuously control two-dimensional target movements independently in the horizontal and vertical directions through ipsilateral and contralateral movements.

Decoding and utilizing dual movements requires consideration of leakage and interference from single movements^42^. Previous findings have been primarily reported at the single-unit level. For example, Rokni et al. documented decorrelation and suppression of ipsilateral neural activity during bimanual reaching in monkeys^58^. Willett et al. conducted a detailed investigation in a paralyzed patient with an implanted MEA and found that, during dual movements, neural representations of the primary effector remained largely stable, whereas those of the secondary effector were relatively diminished^33^. Furthermore, fMRI studies of passive flexion of single or multiple fingers have revealed neural activity patterns specific to finger combinations^59^, suggesting that dual-movement encoding is unique. More recently, iBCI studies of multi-finger gestures summarized and proposed the pseudo-linear summation view of dual movements^43^. However, due to the volume conduction effect of the dura, dual-movement analyses in the eECoG space must also rule out the possibility of simple potential summation. Our study provides evidence at the level of epidural field potentials for the first time, showing that in the HG spatio-spectral feature space, dual movements are closer to their constituent single movements, with the primary effector contributing more and contralateral movements suppressing ipsilateral ones. For ipsilateral dual effectors, such a synergistic pattern may suggest that when neural populations controlling different effectors are simultaneously active, interactions occur. This mechanism can be simulated through a recurrent neural network with static input drive of unequal magnitudes^43^. For homologous bilateral dual movements (eg., left and right hand dual movements), this may instead imply local inhibition mediated by callosal projections^58^. From a neural decoding perspective, this property suggests that treating dual movements as specific categories and mapping them between their two constituent single movements is advantageous for improving information transfer rates, as the impact of interference is minimized.

Another contribution of this study is the introduction of a theoretical model and optimization method for arbitrary mapping paradigms in motor BCI. In prior research, mapping rules for movements were typically designed in an arbitrary manner. More recently, attempts have been made to optimize mapping schemes by using generative models of iBCI signals to allow healthy participants to artificially experience different two-dimensional control strategies^60^. In contrast, we established a BCI channel model capable of offline simulation of two-dimensional directional control, offering a fine-grained optimization approach for mapping vector design as well as quantitative offline estimation. We anticipate that the inclusion of more mapping directions and dual movements influences the source entropy, while mapping rules grounded in neural representational structures enhance BCI channel capacity. Indeed, the channel model establishes a “human-in-the-loop” framework^61^, highlighting that achieving the optimal overall information transfer rate requires joint optimization of the task protocol, user intention strategies, signal feature extraction, classification models, movements mapping schemes, and output parameters, rather than optimizing each component independently.

Further development is required to elevate the performance of this BCI system and to extend its application. First, increasing the number of eECoG electrodes and designing higher-density grids would allow signal acquisition to cover a wider range of motor-tuned cortical areas, thereby improving spatial resolution and enabling the decoding of fine-grained motor movements, such as complex hand gestures. Second, achieving multi-degree-of-freedom control benefits from incorporating a larger repertoire of distinguishable movements into the paradigm, which typically requires extensive pre-screening by experimenters. Designing joint learning frameworks that encourage participants to spontaneously generate separable neural patterns^30^ could enable a more scalable and personalized approach. Finally, implementing automated calibration routines^62,63^ would facilitate long-term use; given the stability of eECoG signals, such calibration is likely to be easier to deploy and more reliable compared with microelectrode-based BCIs. In summary, wireless minimally invasive BCI solutions hold promise for enabling home use human computer interaction and assistance for patients with motor impairments caused by neurological disorders.

## Data Availability

All data produced in the present study are available upon reasonable request to the corresponding author.

## Acknowledgement

We would like to express our gratitude for the commitment and trust of our volunteers and their families. We also thank Shunchang Ma, Jiang Li from Beijing Tiantan Hospital, Capital Medical University; Yujing Wang, Xiaoshan Huang, Yunjing Li and Mengjie Chen from Neuracle Technology Co., Ltd.; and Ze’ao Xiong, Le Yu, Xingrui Chen, and Wen Zhu from Tsinghua University for their assistance with this work.

## Method

### Clinical-trial overview and the participant

The participant in this study (identified as TT01) provided informed consent and was enrolled in the NEO brain-computer interface clinical trial (clinicaltrials.gov Identifier: NCT05920174). The study was approved by the Ethics Committee of Beijing Tiantan Hospital, Capital Medical University. TT01 granted permission for the publication of photographs and videos containing his likeness. All procedures were conducted in accordance with relevant guidelines and regulations.

Participant TT01 is a male (at the time of data collection) who had sustained tetraplegia due to an accident, diagnosed with C4 complete spinal cord injury (AIS-A). A. The implantation surgery of the NEO BCI was performed in Beijing Tiantan Hospital. TT01 retained only very limited voluntary motor function: slight flexion of the left elbow, some visible movements of the face and shoulders, and partial strength in the right biceps, though insufficient to overcome gravity.

### Functional MRI Localization

Before the implantation surgery, the patient underwent head MRI on a 3.0 T scanner (Siemens Magnetom Prisma) to localize brain regions associated with hand sensory and motor functions. Functional MRI (fMRI) was acquired using a gradient-echo echo-planar imaging (EPI) sequence with the following parameters: matrix size 64 × 64, voxel resolution 3.5 × 3.5 × 3.5 mm^3^, repetition time (TR) 2000 ms, echo time (TE) 30 ms, and 36 axial slices covering the whole brain. In addition, to enable geometric distortion correction, a set of EPI images with reversed phase encoding (posterior–anterior direction, 11 volumes) was acquired. A resting-state fMRI scan consisting of 300 volumes was also performed. For structural imaging, a T1-weighted magnetization-prepared rapid gradient echo (MPRAGE) sequence was acquired with the following parameters: matrix size 224 × 224, voxel resolution 1 × 1 × 1 mm^3^, and 192 sagittal slices.

In this study, a functional magnetic resonance imaging (fMRI) paradigm was employed to localize the participant’s sensorimotor regions associated with hand movement and somatosensory processing. The paradigm comprised two tasks designed to assess afferent sensory processing (ascending pathways) and motor control (descending pathways). The first task was a passive movement condition, during which two experimenters assisted the participant in performing passive fist-clenching movements of the left and right hands. This task was intended to stimulate the sensorimotor pathways and elicit activation in the hand-related somatosensory regions. The second task was a motor imagery condition, requiring the participant to imagine performing voluntary hand grasping with either the left or right hand, thereby engaging motor intention–related brain areas. The motor imagery task consisted of two conditions. In the movement condition, the participant was presented with an arrow pointing to either the left or right hand, accompanied by the textual cue “Begin imagining left/right hand movement.” In the rest condition, the participant was shown an image of a teacup along with the textual instruction “Rest, relax, and stay rest.” Both conditions lasted 14 seconds, with left- and right-hand imagery alternating in sequence, repeated for a total of 14 cycles.

### Neurosurgical Operation Planning

Based on the MRI, CT, and fMRI activation results, we developed a surgical implantation plan. The target areas for electrode implantation were the precentral gyrus and the postcentral gyrus, with 4 channels planned for each area. To ensure accurate electrode placement within the fMRI activation area (Extended Data Fig. 2c), we performed 3D modeling using imaging data. The implantable pulse generator was positioned within the flat portion of the skull posterior to the ear, with considerations including avoidance of the temporalis muscle and ensuring sufficient skull thickness for bone-bed preparation. To obtain an accurate topographic map of skull thickness, we developed an algorithm based on CT images to calculate and visualize the skull thickness. During the extraction of the geometric structures of the inner and outer surfaces of the skull, we adopted the SwissSkullStripper algorithm^64^. To calculate the distance between the points on the outer and inner surfaces of the skull and thus obtain the skull thickness, we used the nearest neighbor method. To improve the robustness of the calculation, the number of nearest neighbors in this study was set to 20, corresponding to a resolution of approximately 1.62 mm on the skull surface. Given that the point cloud data of the inner and outer surfaces of the skull were sparse, low-dimensional, but large in volume, we chose to use the K-Dimensional tree (KD tree) method to construct a binary tree structure for the data, thereby accelerating the calculation process. Finally, we assigned the skull thickness information as a color attribute to the point cloud map of the outer surface of the skull and achieved visualization with the help of blender software, providing guidance for the implantation location of the internal device (Extended Data Fig. 2d).

### Experimental Design and Session Structure

In this study, all experiments and data collection were conducted between 36 days post-implant and 523 days post-implant, spanning a total of 225 scheduled days. On each scheduled day, we carried out approximately 1-3 hours of experiments, which included multiple 6-13-minute data collection sessions or test sessions. During the experiments, the participant was seated in a wheelchair with back and leg support. A computer monitor positioned in front of the participant displayed text and images to indicate which movement to perform and when to perform it. The programs controlling the data collection sessions were developed in Python using the PsychoPy library, while the programs for online brain computer interface control experiments were developed in Python and MATLAB.

In this study, the neural activity analyses were primarily based on data collected across 35 sessions conducted between postoperative day 104 and day 232. All sessions followed a delayed-instruction paradigm (Extended Data Fig. 3a). TT01 was instructed to attempt, but not execute, specific movements. The eight movement conditions consisted of single-joint actions (“left elbow flexion, left hand grasp, right elbow flexion, right hand grasp”) and paired-joint actions (“left elbow & left hand, left elbow & right elbow, left hand & right hand, right hand & right elbow”). Each session comprised four data-collection blocks. At the beginning of each block, a textual cue indicated the movement type for the subsequent trials. Each block contained 20 consecutive trials of the same movement. a textual instruction appeared at the center of the screen during the delay phase, prompting TT01 to prepare for the designated movement. The delay duration was drawn from a uniform distribution between 1.4 and 1.8 seconds. At the onset of the go cue (a specific image displayed at the center of the screen), TT01 immediately attempted the movement and sustained the attempt or imagery for 2.5 seconds. This was followed by a rest phase, indicated by a screen icon, during which TT01 relaxed and remained at rest for 2.0 seconds.

### Neural Data Processing

Epidural neural signals recorded by the NEO brain–computer interface were preprocessed using the MNE-Python toolbox. The signals were high-pass filtered at 1 Hz and notch-filtered at 50, 100, and 150 Hz (transition bandwidth: 3 Hz). A common average reference was then applied by subtracting the mean signal across the electrode array from each channel.

To evaluate spectral modulations during movement execution, we computed time–frequency representations (TFRs) using the discrete prolate spheroidal sequences (DPSS) taper method, and further derived event-related spectral perturbations (Extended Data Fig. 3b). For the calculation of root mean square (RMS) amplitude (Fig. 1d), power spectral density was estimated using Welch’s method, followed by square root transformation.

For the online BCI task, neural signals were processed in 1 second sliding windows with a step size of 200 ms. A second-order Butterworth high-pass filter (1 Hz cutoff) was applied in real time, followed by common average referencing.

### Dual movement neural pattern

To examine the separability of neural signals across the eight movement conditions, we computed the spatiotemporal patterns of RMS amplitude (Extended Data Fig. 4a) for each movement (subtracting the resting baseline from -1000ms to 0ms for each trial), and concatenated the channels (8 channels in total) with the frequency points (*fb*=76 uniformly sampled frequency points between 0-150Hz). We used the Mahalanobis distance to evaluate the pairwise statistical distance *D*_*ij*_ between the neural centroids of RMS amplitude spatio-spectral patterns (assumed to follow a multivariate Gaussian distribution) across all eight movements.

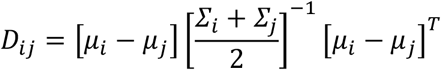

where μ_*i*_ ∈ ℝ^1×(*ch*×*fb*)^ is the conditional average spatio-spectral pattern vector of movement *i* and ℝ^(*ch*×*fb*)×(*ch*×*fb*)^ is the covariance matrix of the spatio-spectral patterns of multiple trials of movement *i*. That is, the Mahalanobis distance represents the pairwise distance between the patterns of any two movements, scaled by the pooled multivariate variance. To reveal the similarity structure among the spatio-spectral pattern distributions of different movements, we performed hierarchical clustering on the average Mahalanobis distance matrix using Ward’s criterion^65^, and visualized the results with a scaled dendrogram (Fig. 2b).

### Low-dimensional projection of eight movements

We aimed to identify the principal neural axes that constitute a shared subspace across different movements. To this end, we projected the high-dimensional spatio-spectral patterns of onto a lower-dimensional space, thereby providing an intuitive visualization of the representational similarity of the eight movements within the neural subspace. Our approach was mathematically inspired by the classical demixed-principal component analysis (dPCA) method^66^. Specifically, we formulated a generative assumption for the neural feature data, in which the pattern vector ***X*** of a single trial can be expressed as the sum of the task-averaged component 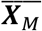 and the noise term ***X***_*noise*_. Unlike standard dPCA, here the neural patterns are defined at a single time point (time dimension equal to 1), and only one marginalization condition— “movement” (***M***)—is considered.

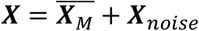

Suppose there are *k* trials for each of the *m* movement conditions, then the size of the matrix ***X*** in the above formula is (*hc* × *f*)*b* × (*k*× *m*). 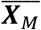 are also matrices of (*ch* × *f*)*b* × (*k* × *m*) where (*hc* × *f*)*b* × *m* different values (average under each condition) are replicated *k* times and corresponding to the index of ***X***. Such a decomposition provides an interpretable and comprehensive structural overview, as it represents the variance related to specific tasks (movements) in the experiment. The loss function of the decomposition is defined as

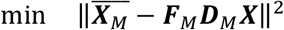

where ***F***_***M***_ is the encoder matrix of *q* columns, that is, the main neural axes related to movements we want to find, and ***D***_***M***_ is the decoder matrix of rows.

For the single movements neural axes and dimension reduction analysis in Fig. 2a, we calculated the standardized HG frequency band (50-150Hz) RMS amplitude spatio-temporal features of single/dual movements (subtracting the average resting baseline under all conditions). For single movements, we obtained the HG band pattern ***X***_*sing*_ and the task-average matrix 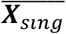. The minimization problem for the above loss function can be formulated as a reduced-rank regression problem, which was solved by first computing an ordinary least squares solution matrix ***A***_*OLS*_.

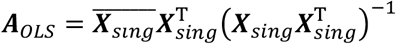

We performed PCA on ***A***_*OLS*_***X*** and take the first *q* eigenvectors (here *q* = 2) to obtain the optimal low-rank approximation:

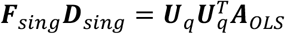

where ***U***_*q*_ ∈ ℝ^(*ch*×*fb*)×*q*^ are the first *q* principal components (left singular vectors) of ***A***_*OLS*_***X***. Finally, we got encoder matrix ***F***_*sing*_ = ***U***_*q*_ and decoder matrix 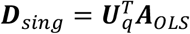. We used ***F***_*sing*_ to reduce the dimension of ***X***_*sing*_ to obtain a two-dimensional projection 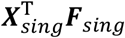 of the single movements’ subspace.

For the dual movements dimension reduction in Fig. 2c, we project the dual movements into the single movement subspace to inquire about the representational structure of single/ dual movements. We use ***F***_*sing*_ to reduce the dimension of ***X***_*dual*_ and obtain the two-dimensional projection 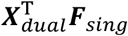 of the dual movements.

### Dual movement linear regression analysis

We use linear regression analysis to verify which of the three predefined hypotheses the neural coding is more consistent with when the two effectors move simultaneously. Considering that the energy in the HG (50-150Hz) frequency band is a proxy for the local neuronal firing activity, our analysis is focused on the HG frequency band. For each dual movement (taking the simultaneous movement of effector *a* and effector *b* as an example), we establish a linear regression model for the spatio-temporal pattern *X*_*a*&*b*_ (the feature dimension is *ch* × *bf*). We select different sets of basic features as explanatory variables.

*Self-basis*:

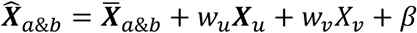

*Orthogonal-basis*:

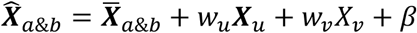

*Component-basis*:

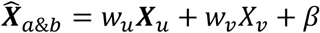

Where *X*_*u*_ ∈ ℝ^1×(*ch*×*f*_*HG*)^ and *X*_*v*_ ∈ ℝ^1×(*ch*×*f*_*HG*)^ are two specific bases, *w*_*u*_ and *w*_*v*_ are their corresponding regression coefficients, *β* is the bias term. We designed three sets of bases for the linear regression model. For *self-basis*, we calculated the first two principal components *X*_*a*&*b*_*pc*1_ and *X*_*a*&*b*_*pc*2_ of *X*_*a*&*b*_. It provides the theoretically highest fitting effect that two sets of bases can achieve for *X*_*a*&*b*_. For the *orthogonal-basis*, the last two principal components 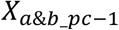 and 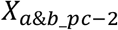 are taken as the bases. Since these two sets of bases are orthogonal to the first two principal components, it can represent the situation of the *Outside manifold*, and it’s expected that will not fit well. For *component-basis*, we constructed four groups of bases, each group consisting of the conditional average patterns of two single movements (LE-LH, LE-RE, LH-RH, RE-RH). Each of these four groups of bases spans a subspace fully supported by single movement information. We also constructed the *non-component-basis* that is completely non-corresponding to specific dual movement (RE-RH regressed on LE&LH, LH-RH regressed on LE&RE, LE-RE regressed on LH&RH, LE-LH regressed on RE&RH). We evaluated different hypotheses using the goodness of fit *R*^2^ of the regression model:

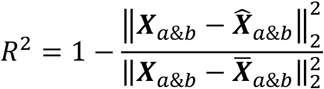

To more rigorously verify whether the dual movements is merely a sensor space superposition of the neural activities of single movements, we constructed surrogate dual movements data by summing single movements signals and conducted the same linear regression analysis. Denote the LPF signal (distributed source hypothesis) of the subdural sensorimotor cortex as ***z***, and the field potential signal recorded by the epidural sensor as ***s***. Under the *Observation space superposition* hypothesis, the surrogate dual movements signals 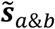 should be:

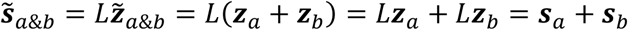

where is the leadfield matrix from the LPF of the cortex to the eECoG electrodes. Therefore, we generated surrogate dual-movement neural signals 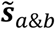 by randomly pairing and summing neural signals from two different single movement trials (***s***_*a*_, ***s***_*b*_), while keeping the number of surrogate trials equal to that of the actual dual-movement dataset. After applying common average referencing, we computed the power spectral density (PSD). Given that two randomly paired single movement signals can be considered approximately independent, the direct summation will amplify signal power. Moreover, previous studies have reported that dual movement neural activity exhibits normalization. Therefore, we multiplied the PSD by a normalization factor to ensure that the surrogate and actual dual-movement RMS amplitudes were on the same order of magnitude

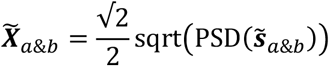

### Movement Decoding and Target Control

For BCI two-dimensional target control, we first selected the BCI movement repertoire *M*, eight movements in total. The linear discriminant classifier was used to classify different types of movements (movements in *M* plus rest). Neural data were segmented (segment length: 1 s) to construct the training set. For each trial (with movement onset at time 0), data from the [−2s, −1s] interval were labeled as rest, while data from [0.1s, 1.1s] and [1.1s, 2.1s] intervals were labeled according to the corresponding movement. We applied a linear discriminant classifier to the 8-channel standardized spatio-spectral features (logarithmic PSD from 0–100 Hz computed via Welch’s method). We assumed that the *X* follow a multivariate Gaussian distribution with a shared covariance matrix *Σ* across different classes.

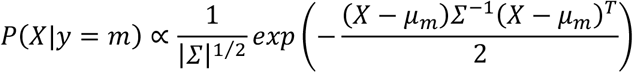

The log posterior probability is obtained by the Bayes’ formula.

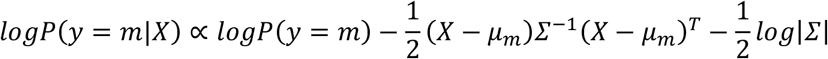

Here, we assumed equal prior probabilities *P*(*y* = *m*) for all classes. For each sample, the logarithm of the posterior probabilities was first shifted by subtracting the maximum value across all classes, then exponentiated and normalized to yield the estimated posterior probability *P*(*y* = *m*|*X*). Next, we used a set of mapping vectors 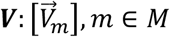, assigning a “preferred direction” 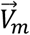 to each movement. The posterior probabilities were used to compute a weighted sum over the “preferred directions”. Additionally, we designed a rest-gating factor *σ*_*rest*_ = 1 − *P*(*y* = |*X*)^¼^ to ensure that the velocity approaches zero when the rest probability is high (i.e., near the rest state). This yielded the final velocity.

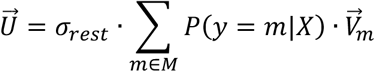

For the selection of the mapping vector set ***V***, unless otherwise specified, we use a fine-grained Sing.&Dual 8D vector set (Extended Data Fig. 7a top). It is obtained by optimizing the mutual information between the source and the sink of the two-dimensional control BCI channel (see Method “Optimization of two-dimensional Control Mapping Vector Set”).In the subsequent analysis, we also compare two other representative vector sets. The Sing. Movement 4D vector set (Extended Data Fig. 7a left) only depends on four movements: LE, LH, RE, and RH.

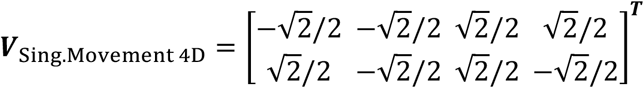

The Sing.&Dual 4D vector set (Extended Data Fig. 7a right) is grouped into 4 groups according to {LE, LE&RE}, {LH, LE&LH, LH&RH}, {RE}, {RH, RE&RH}, and then corresponds to

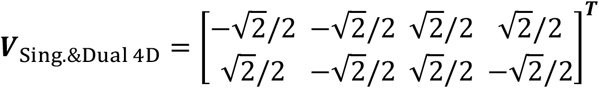

### Center-out task experiment

In the online Center-out task (Extended Data Fig. 6a), the subjects were required to perform various attempted movements to control a controlled target on the screen to touch the cued target around the screen using a brain-computer interface. The radius of the controlled target is 0.05 units, and its initial position is at the center of the screen. The maximum distance that its center can be away from the center of the screen is 0.41. The radius of the cued target is 0.06, and it may appear at one of the 8 equally spaced (45 degrees) positions around the screen (the center of the circle is 0.40 away from the center of the screen).

Each session of the online Center-out task contains 3 rounds. In each round, 8 different cued targets in random order will be traversed (i.e., a total of 24 ball-touching trials). For each touch task, the cued target is displayed on the screen, and the controlled ball appears at the center of the screen (but its speed is fixed at 0 for 1 second to provide the subject with preparation time). After 1 second, the subject can normally control the controlled ball, and its position trajectory is

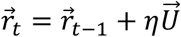

where η =0.025. The subject has a maximum of 8 seconds to drive the controlled sphere to touch the cued target (if the distance between the centers of the two spheres is less than or equal to 0.11 units, it is judged as a hit). Either a hit or a time-out will lead to the next touch task.

The evaluation the Center-out task using the Sing.&Dual 8D vector sets in the first month (denoted as month 0) included 5 sessions. After that, 3 sessions were conducted on a specific day each month to track long-term performance. The tests for the Center-out task using the Sing. Movement 4D and Sing.&Dual 4D vector sets were carried out in month 1, each containing 3 sessions. Before the tests, we usually conduct 10-20 minutes of practice to help the subjects recall and get familiar with the operation.

### Web-grid Task Experiment

In the online web-grid task (Extended Data Fig. 6b), participants were required to perform various trial movements to use a brain-computer interface to control a controlled ball on the screen to touch the cued target on an 8×8 grid (each square has a size of 0.1 unit). The radius of the controlled ball is 0.04 units, and its initial position is at the center of the screen, with its movement range restricted to the grid. The radius of the cued target is 0.05 units.

Online web-grid evaluation uses Sing.&Dual 8D vector sets. The test in the first month (in month 1) consists of 5 sessions, and thereafter, 3 sessions are conducted on a specific day of each month to track long-term performance. Each session contains 24 trials. After the start of each trial, the cued target may be at the center of any one of the 64 squares. The subject then controls the controlled ball to move from its current position. The subject has a maximum of 14 seconds to complete a hit (a hit is determined when the distance between the centers of the two balls is less than or equal to 0.08 units). If a hit is made in advance or the time limit is exceeded, it will jump to the next trial. The rest of the parameter settings, control methods, etc. of the web-grid evaluation are the same as those of the Center-out task.

### Two-dimensional Control Evaluation Metrics

We use metrics such as hit rate, information transfer rate, and path distance ratio to evaluate the performance of subjects in performing the center-out task.

1. The hit rate *p* is defined as the ratio of the number of trials in which the touch is completed within the time *T*_*max*_ to the total number of trials.
2. Fitts’ ITR. It is used to measure the information transfer rate in continuous two-dimensional control tasks^67^. For a target with size (diameter) *G*, if the distance to the target is *D*, there is

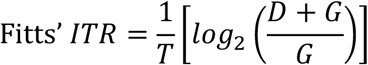

where *T* is the time cost of one touch (trials that are not successful within *T*_*max*_ are not counted). In this study, the size *G* is equivalent to twice the minimum distance determined as a hit.
3. The Trajectory distance ratio is defined as the ratio of the trajectory path of the center of the controlled ball in a successful touch trial to the theoretically shortest path.
4. Trajectory error is defined as the integral of the error by which the actual path deviates from the shortest path^68^.
5. Trajectory variability is defined as the variance of the error of the actual path deviating from the shortest path^68^. A larger Trajectory variability indicates that the path is more tortuous.
6. Hitting ITR. Considering that the hit rate and hit time are not consistent in some cases (e.g., control is only efficient in certain directions), we define Hitting ITR to comprehensively measure the two-dimensional control performance. Referring to the ITR definition in BCI multi-class classification, for a task with *N* cued targets (in this study, *N* = 8)

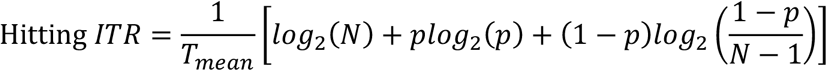

Where *T*_*mean*_ is the average time required to hit the target (if it exceeds *T*_*max*_, then take *T*_*max*_).

### Offline Simulation of Center-out Task

For the offline simulation of the Center-out task, we draw on the subject model from the macaque BCI research, assuming that at each moment the subject intends to move the controlled target toward the cued target^62^. Accordingly, the target velocity vector at the current time point, *S* ∈ ℝ^1×2^, was defined as the vector pointing from the current controlled target position to the cued target position (Extended Data Fig. 7b, bottom), while the output Y ∈ ℝ^1×2^ corresponded to the controlled target velocity (scaled by a proportional factor).

We decompose Center-out into a two-dimensional velocity control task and its integration process. Assume that the BCI repertoire provides a total of *L* protocol movements. The simulation process will generate the controlled velocity vector Y ∈ ℝ^1×2^ based on the target velocity vector *S* ∈ ℝ^1×2^, the movement classification confusion matrix ***C*** ∈ ℝ^*L*×*L*^, and the mapping vector set 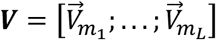. We constructed a virtual subject model, which can generate the probability distribution ***P***_*V*_(***M***|*S*) ∈ ℝ^1×*L*^ of the movement intention ***M*** based on *S*. Denote *α* and *β* as the two movements whose vector 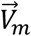 re closest to *S*, with *S* lying between 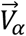 and 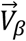 (Extended Data Fig. 7b, top). Define

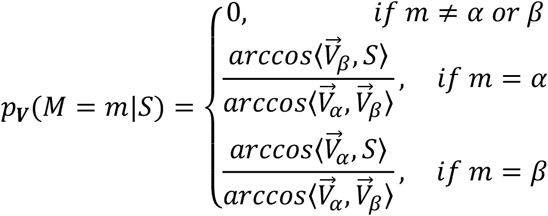

The virtual subject receives the visual input *S* (information source) and generates the intention *m∼****P***_***V***_(***M***|*S*). After sampling to get *m*, the movement prediction probability *p*(*y* = *m*|*X*) output by the BCI decoder is the corresponding row ***C***_*m*_ in the normalized confusion matrix, thus obtaining the prediction result 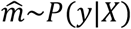. In the simulation, we simplified the two-dimensional velocity control output gating function, let

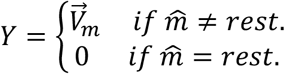

We can obtain that, for a certain target velocity vector *S*

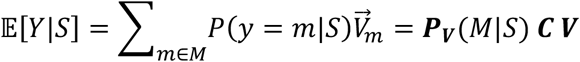

In the simulation, we adopted exactly the same parameter settings as in the real Center-out task (except that the maximum allowed time was changed to 10s). For each set of mapping vectors, we conducted 9 rounds of 8-target traversals, and thus calculated the average hitting time. We also constructed random surrogate mapping vectors sets to evaluate the efficiency of the Sing.&Dual 8D (Extended Data Fig. 7a, top). The eight movements were arbitrarily mapped to eight equally spaced directions 0, π/4, …, 7π/4, yielding a total of 2520 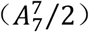 essentially different arrangements (excluding overall rotation or mirror symmetry). We simulated the Center-out task for each surrogate mapping vector and obtained the corresponding average hit time, from which the histogram in Fig. 3f was derived.

### Optimization of two-dimensional Control Mapping Vector Set

Consider two-dimensional velocity control. After observing the controlled target velocity vector *S* (information source), the subject generates an intended movement according to the policy, and the resulting neural activity in the sensorimotor cortex is probabilistically decoded into an estimated movement, which is then mapped to the controlled velocity vector Y. This process can be regarded as a communication channel. Let the directions of the velocity vectors be ϑ_*S*_ and ϑ_*Y*_, and assume 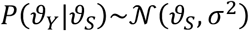. The mutual information between source and sink is

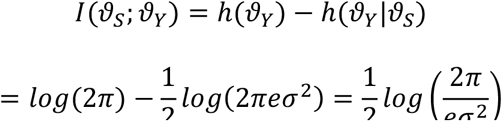

Hance,

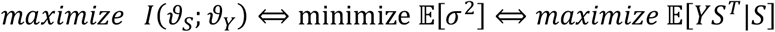

Considering that we derived 𝔼[Y|*S*] = ***P***_***V***_(***M***|*S*) ***C V*** in the previous session, it yields

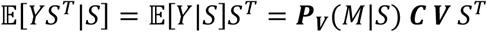

To identify the optimal *V* for full-direction control, target vectors *S* were sampled along the circle at π/36 intervals to construct the matrix *S* ∈ ℝ^72×2^. Using the virtual subject model, we computed the corresponding movement policy matrix ***P*** ∈ ℝ^72×8^ (the subject was trained to follow an 8D vector set ***V***_**0**_ ∈ ℝ^8×2^ with π/4 spacing, which uses the ***V***_Sing.Movement 4D_ as skeleton and maps dual movements to vectors between two single movements, so that here 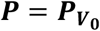. This led to the following optimization problem:

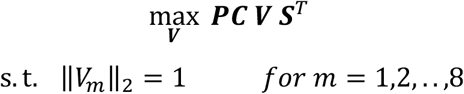

The constraints enforce the magnitude of *V*_*m*_. Applying the method of Lagrange multipliers yields

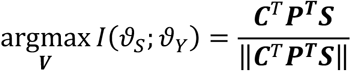

The obtained mapping vector set ***V*** based on the confusion matrix (in Fig. 3b) is shown as Sing.&Dual 8D in Extended Data Fig. 7a. Given the decoder capacity ***C*** and the intended movement policy ***P***, this mapping vector set can theoretically achieve the maximal information transfer rate for two-dimensional velocity control.

### Long-term Stability of Signals and Decoding Performance

To verify the long-term application performance of the bilateral single/dual movement decoding controller, we collected single and dual movement data (one session per day) on 17 scheduled days within 6 months (about 190 days) after the decoding model was trained (denoted as decoder time span Day0, corresponding to the 328th day after implantation). The decoding model was used to classify the single and dual movement trials of each day (Fig. 4b). The data of 4 scheduled days (8 sessions) in decoder time span Day25-100 and Day151-193 were classified to obtain the confusion matrix (Fig. 4a).

We assessed signal stability by monitoring the amplitude and power spectra across different frequency bands during rest (Extended Data Fig. 8a, 8b), covering 60 scheduled days between 28 and 523 days post-implantation. We conducted a long-term daily paradigm in which, at the beginning of each session, the patient maintained a two-minute resting state. The subsequent tasks included 15 paired elbow flexion/rest trials (randomly containing 10 left elbows and 5 right elbows), 15 paired hand grasping/rest trials (randomly containing 10 left hands and 5 right hands), and 15 paired passive left-hand grasping /rest trials. Each trial’s task phase lasted 5 s. During the rest phase, the patient was instructed to remain relaxed for 6 s; the 4–6 s segment was extracted as resting-state data. RMS amplitude was computed, and the average power spectrum in four frequency bands (1–10 Hz, 10–50 Hz, 50–100 Hz, and 100–200 Hz) was calculated using Welch’s method. Averaging across trials yielded the daily signal features.

Epidural electrode contact impedance was measured via milliampere current stimulation and recorded for 280 days spanning the implantation day and the following 18 months (Extended Data Fig. 8c).

---

**Extended Data Figure 1.**
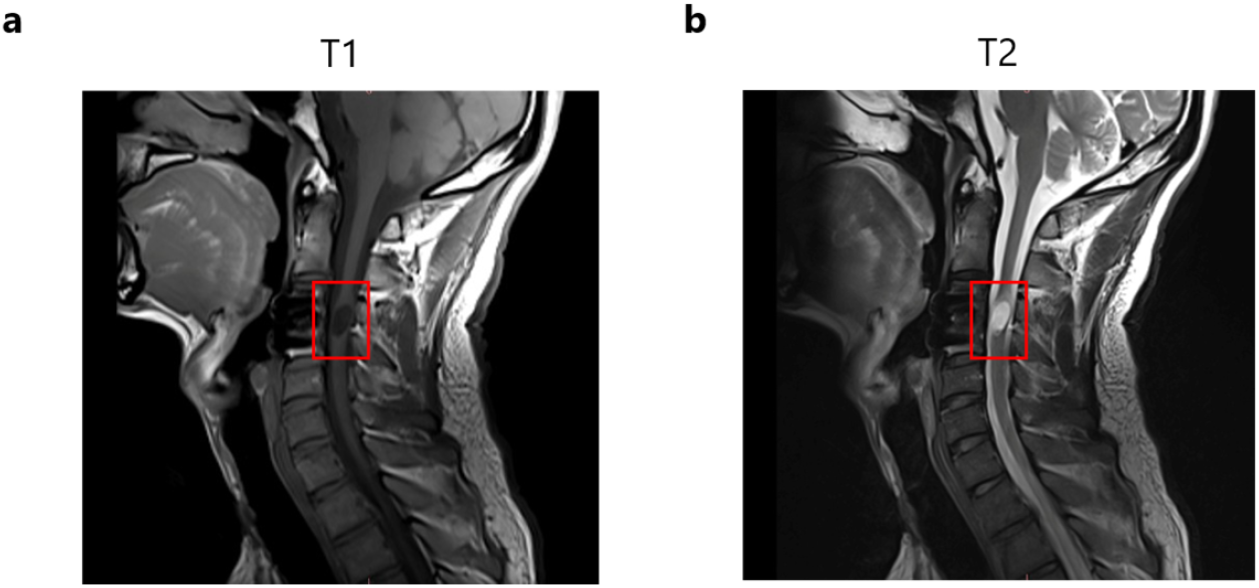
MRI Scans of the Patient’s Head and Neck. **a**. Cervical spine T1 structural image. **b**. Cervical spine T2 structural image; the red box indicates the location of the patient’s spinal cord injury.

**Extended Data Figure 2.**
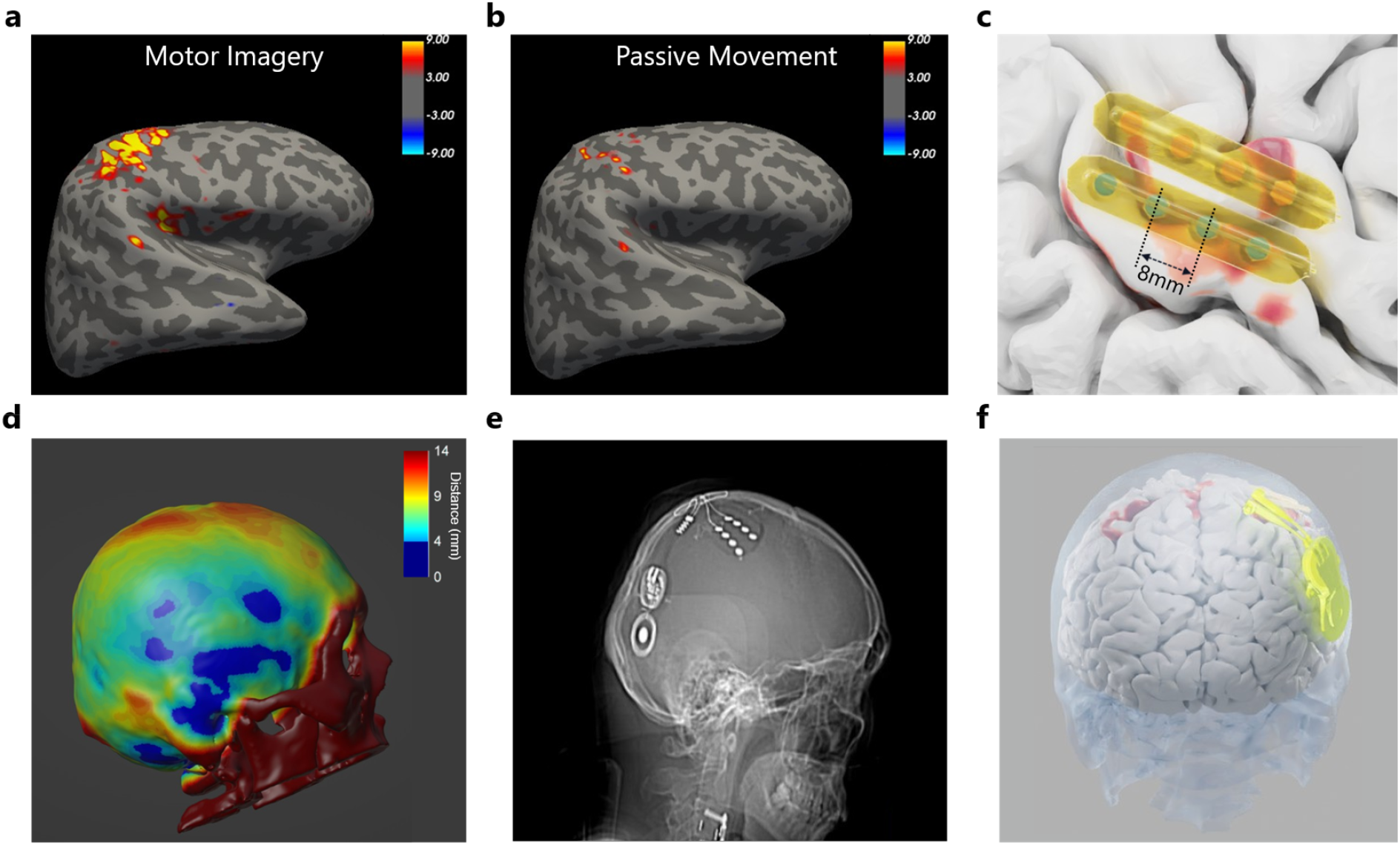
fMRI Localization and Surgical Planning. **a-b**. Functional activation during motor imagery (a) and passive movement (b), with activation values represented as -lg(*p*). **c**. Enlarged view of the diagram of the planned electrodes. Electrodes 1-4 were located on the precentral gyrus and electrodes 5-8 on the postcentral gyrus. **d**. Schematic diagram of the patient’s skull thickness. **e**. Sagittal View of postoperative CT, showing the actual implantation positions. **f**. Rear view of the implantation position of the NEO Brain-Computer Interface

**Extended Data Figure 3.**
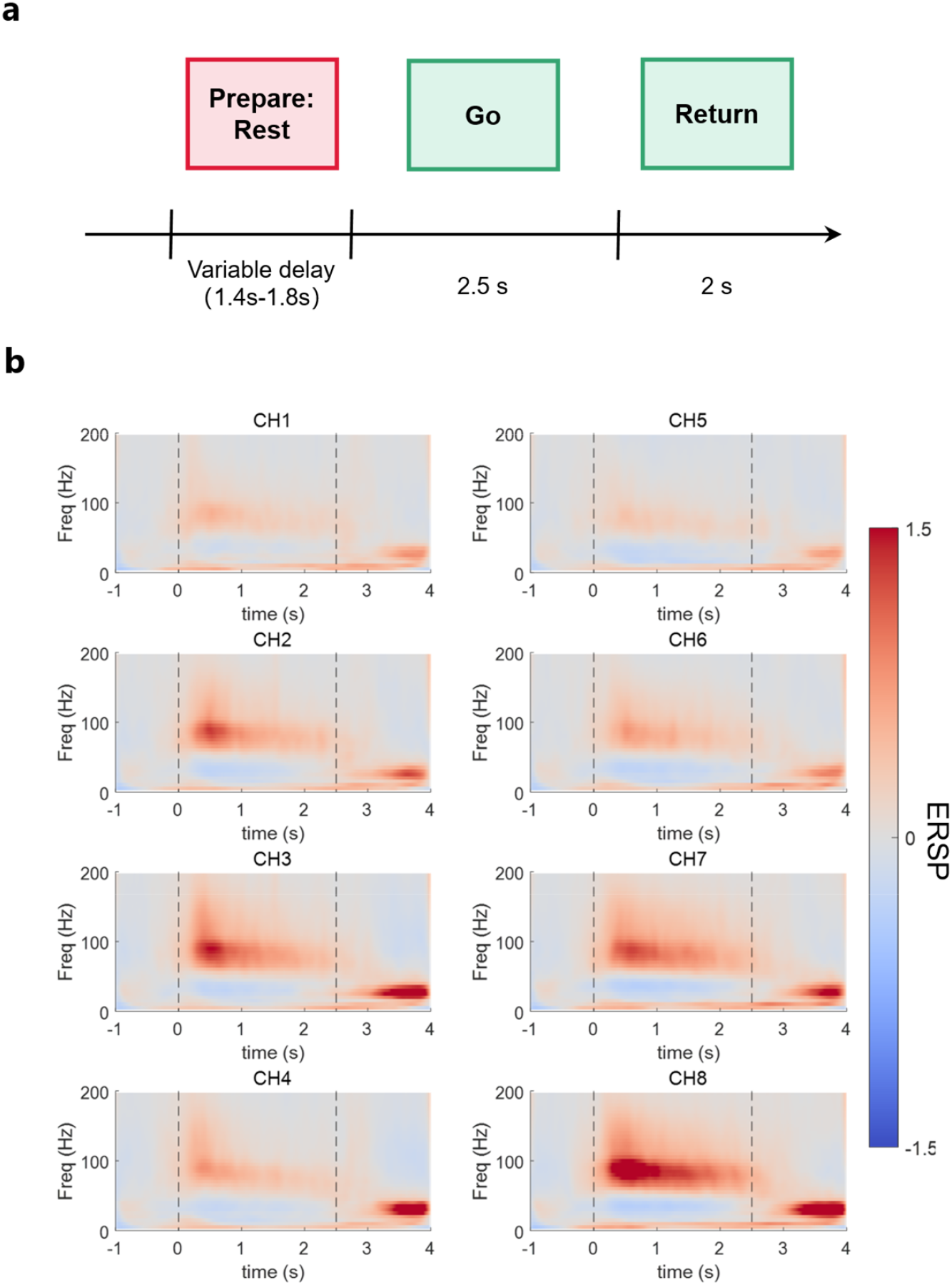
Contralateral and Ipsilateral movements neural coding. **a**. The instructed delay paradigm was used to collect neural data of different ipsilateral and contralateral movements as well as double movements. **b**. The trial-averaged time-frequency response of the attempted contralateral hand grasping movement, presented as Event-related spectral perturbation (ERSP), -1000∼0 ms as the baseline.

**Extended Data Figure 4.**
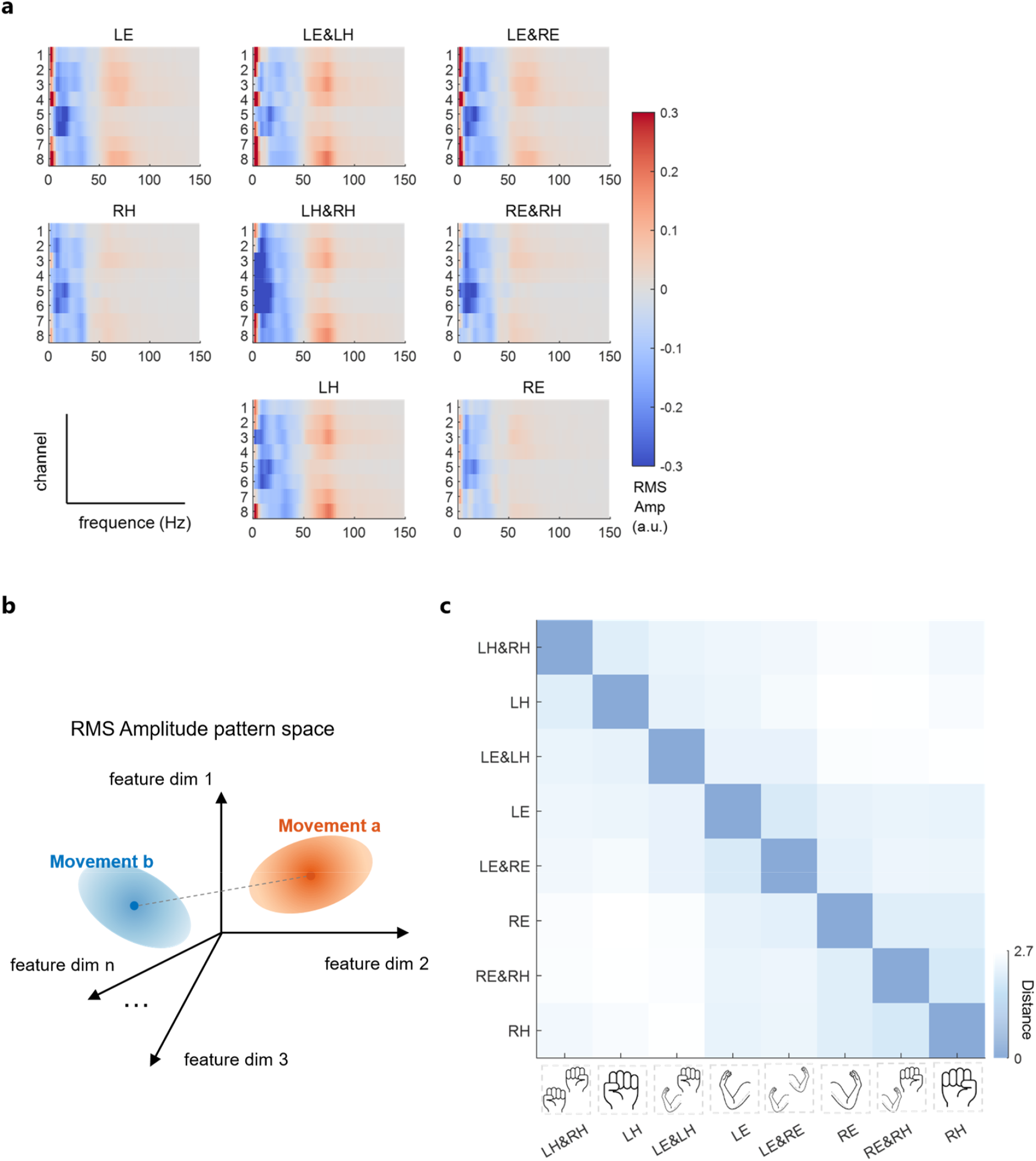
Neural spatio-spectral patterns of single and dual movements. **a**. Average multi-channel RMS amplitude patterns of four single movements and four dual movements. The task phase is from 200∼1200 ms, and the baseline is -1000∼0 ms. **b**. We evaluated the Mahalanobis distance between different movements in the high-dimensional space of the spatio-spectral patterns of the multi-channel RMS amplitude. The neural centroid (the solid circle in the figure above) is the pattern average; the multi-dimensional variance determines the distribution (ellipsoid) around the centroid. **c**. Pairwise Mahalanobis distance matrix between the neural centers of four single movements (left elbow flexion, left hand grasping, right elbow flexion, right hand grasping) and four dual movements (left elbow & left hand, left elbow & right elbow, left elbow & right hand, right elbow & right hand).

**Extended Data Figure 5.**
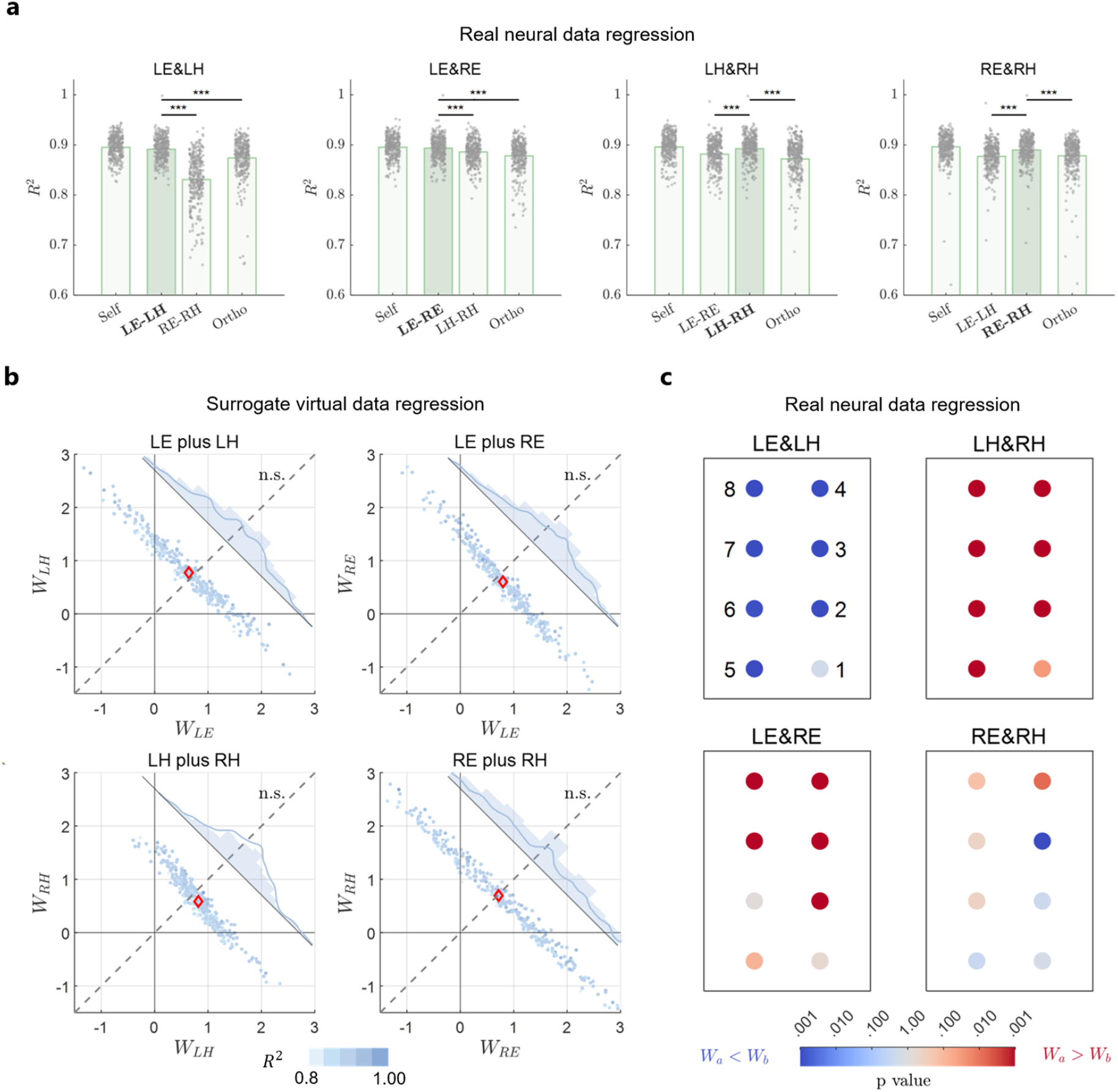
Dual movement regression analysis. **a**. Goodness of fit of dual movement regression under different “bases”. The *self-basis* has the highest goodness of fit. The single movement *component-basis* (average *R*^2^ =0.891 for LE-LH, *R*^2^ =0.893 for LE-RE, *R*^2^ =0.892 for LH&RH, *R*^2^ =0.890 for RE&RH) is significantly greater than (two-sample t-test *p* < 1e-3) the *non-component-basis* (such as LE&LH corresponding to LE-LH, completely non-corresponding to RE-RH), and also greater than the *orthogonal-basis* (two-sample t-test *p* < 1e-2). **b**. To verify that the dual movements is not just a linear superposition in the sensor observation space. We constructed a virtual surrogate dataset by adding the original signals of single movements (n=380 trials randomly paired), calculated its High γ (50-150Hz) RMS Amp. feature, and performed the same regression analysis. For virtual surrogate dataset, the two regression coefficients of the dual movement regression model have no statistically significant difference at the significance level of 0.001 (Wilcoxon signed-rank test *p* = 0.265 for LE&LH, *p* = 0.003 for LE&RE, *p* = 0.022 for LH&RH, *p* = 0.585 for RE&RH). **c**. We carried out the regression analysis electrode by electrode. Colors indicate the significance (paired t-test *p*-value) of the regression coefficient of movement a in the dual movement a&b regression model being different from (marked red if greater, marked blue if less) that of movement b for the corresponding electrode. The regression model basically has good consistency among electrodes.

**Extended Data Figure 6.**
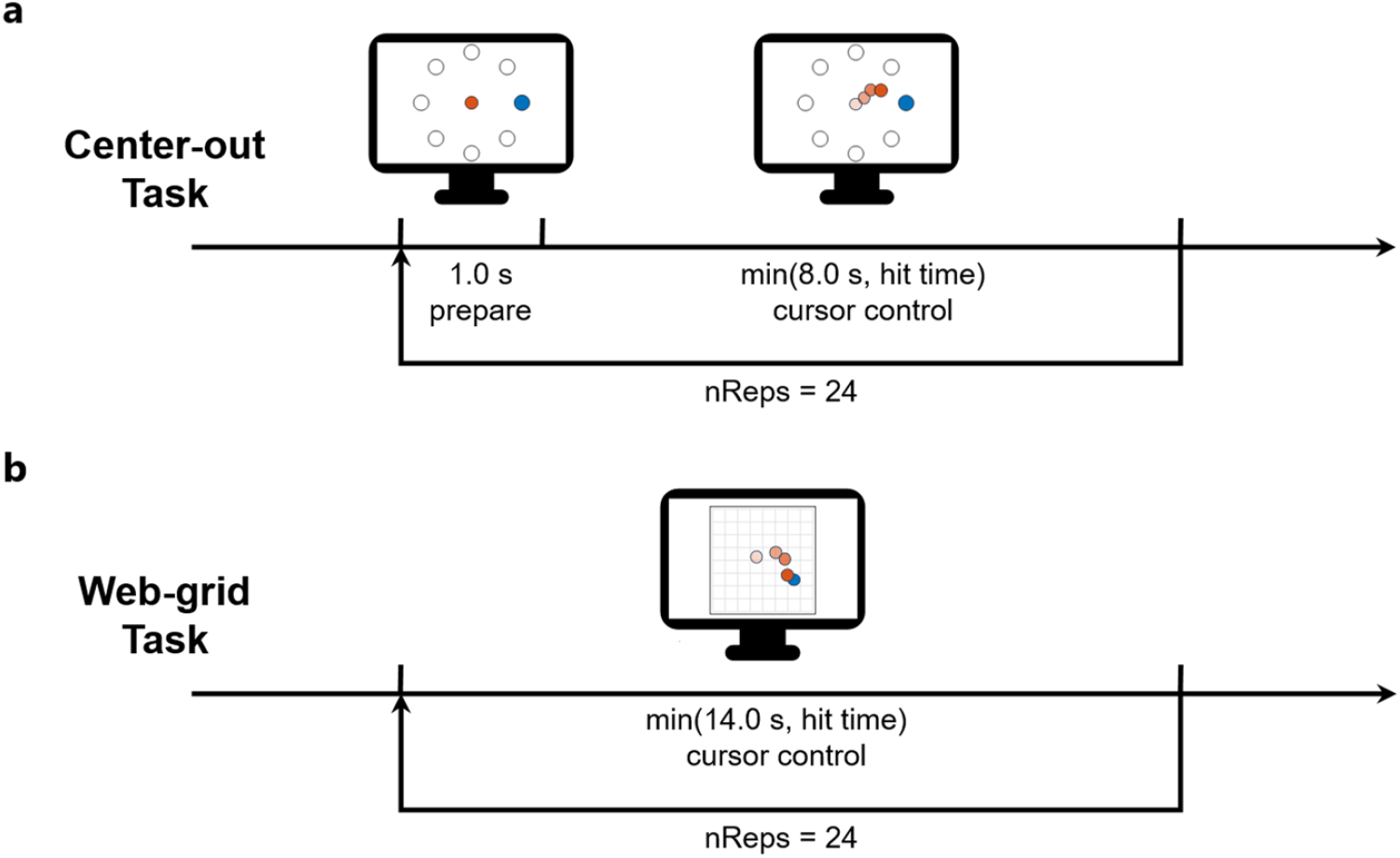
two-dimensional target control paradigm. **a**. Schematic diagram of the Center-out task experimental paradigm (one session). **b**. Schematic diagram of the web-grid task experimental paradigm (one session).

**Extended Data Figure 7.**
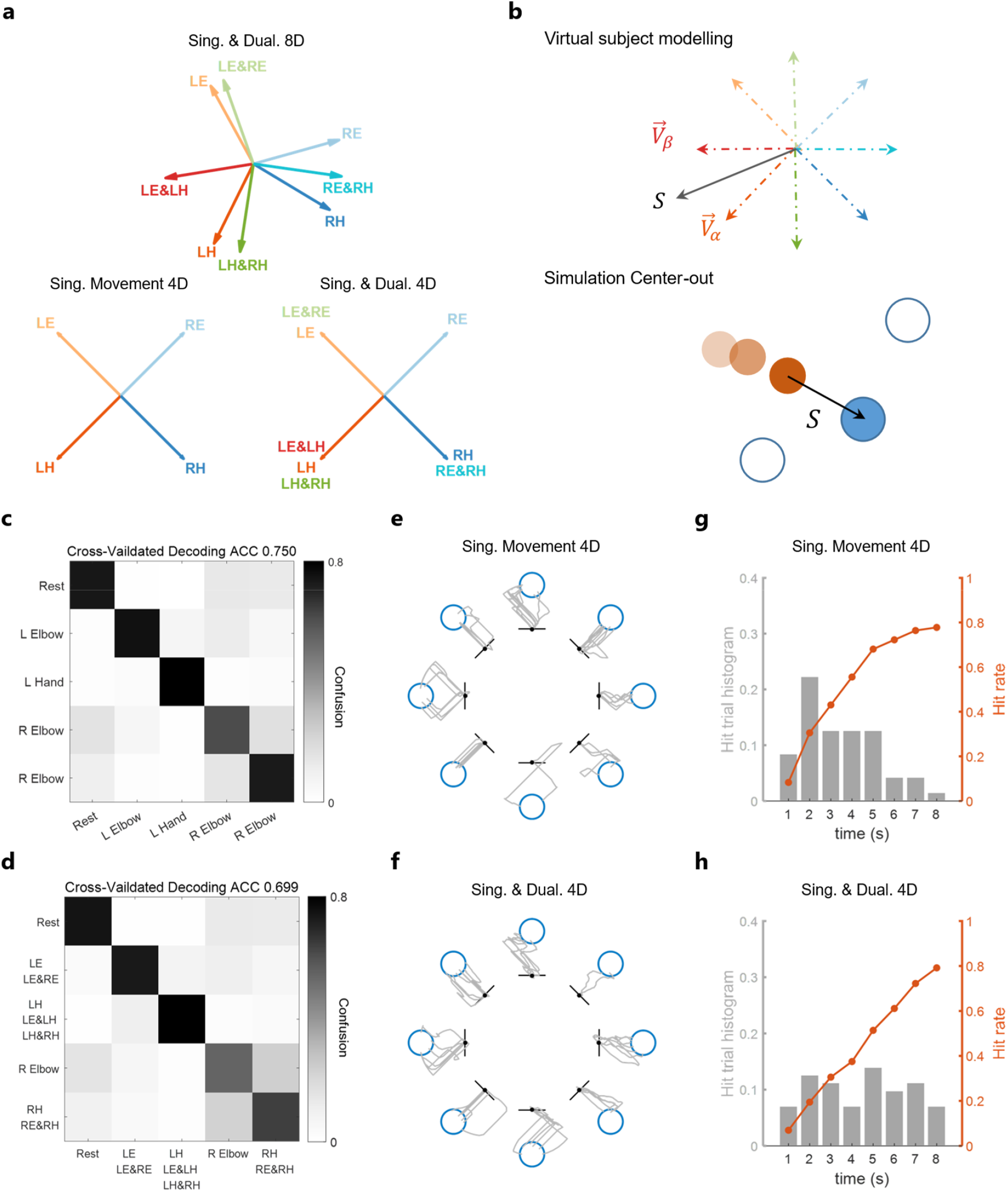
Virtual experiment and mapping vector set testing. **a**. Mapping vector set of Sing. Movement 4D (left), Sing. & Dual. 4D (right), Sing. & Dual. 8D (top). **b**. In offline simulation, the virtual subject generates movement intentions based on the intuitive direction S and the taught mapping vector set (top), where the intuitive direction S (information source) in the Center-out task is the vector from the current position of the controlled target to the cued target (bottom). **c-d**. Movements classification confusion matrices. The top shows four single movements and rest, the bottom shows four grouped movements and rest. Obtained by averaging 100 ten-fold cross-validations. **e-f**. Online Center-out task trajectories of the Sing. Movement 4D mapping vector set (top) and the Sing. & Dual. 4D mapping vector set (bottom). **g-h**. Histograms of hit time in the online Center-out task for the Sing. Movement 4D mapping vector set (top) and the Sing. & Dual. 4D mapping vector set (bottom), and the hit rate within a specific time.

**Extended Data Figure 8.**
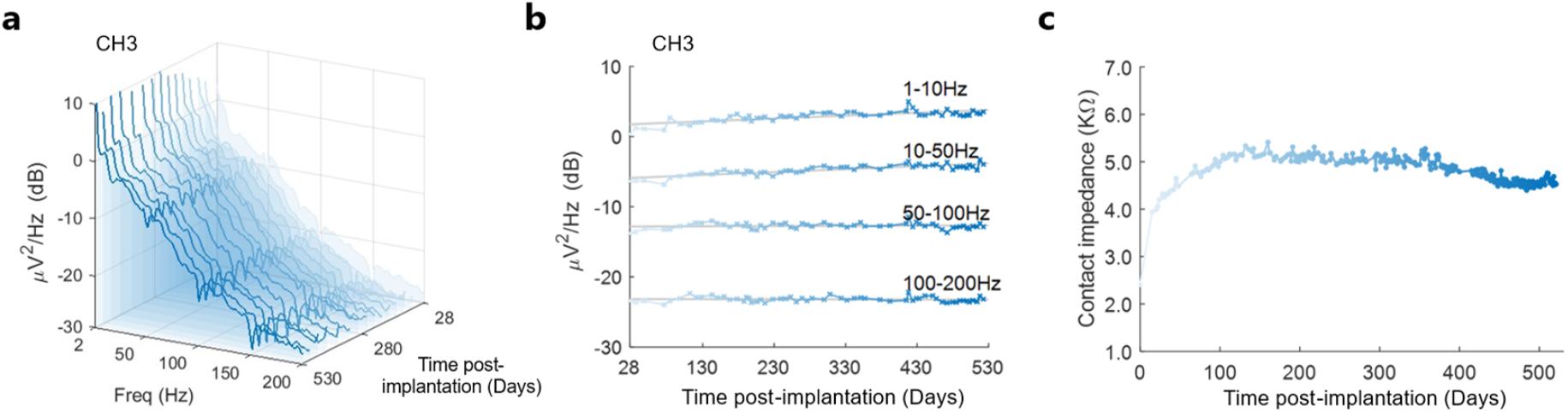
Long-term stability analysis. **a**. Regularly obtain resting-state recordings (continuous recordings for up to 18 months) to evaluate the change in signal quality over time. The power spectral density of the ECoG signal (from CH3) shows long-term signal stability. **b**. The power spectrum of the eECoG signal sub-bands measured by a typical electrode (from CH3). The increase in signal power is limited, with an average of 4.1e-3 dB/day in the 0-10 Hz band, 4.2e-3 dB/day in the 10-50 Hz band, and no significant change in the 50-200 Hz band.**c**. The contact impedance averaged over 8 channels initially increases and then remains stable.

